# Clinical validation of colorimetric RT-LAMP, a fast, highly sensitive and specific COVID-19 molecular diagnostic tool that is robust to detect SARS-CoV-2 variants of concern

**DOI:** 10.1101/2021.05.26.21257488

**Authors:** Pedro A. Alves, de Ellen G. Oliveira, Ana Paula M. Franco-Luiz, Letícia T. Almeida, Amanda B. Gonçalves, Iara A. Borges, Flávia de S. Rocha, Raissa P. Rocha, Matheus F. Bezerra, Pâmella Miranda, Flávio D. Capanema, Henrique R. Martins, Gerald Weber, Santuza M. R. Teixeira, Gabriel Luz Wallau, Rubens L. do Monte-Neto

## Abstract

The COVID-19 pandemics unfolded due to the widespread SARS-CoV-2 transmission reinforced the urgent need for affordable molecular diagnostic alternative methods for massive testing screening. We present the clinical validation of a pH-dependent colorimetric RT-LAMP (reverse transcription loop-mediated isothermal amplification) for SARS-CoV-2 detection. The method revealed a limit of detection of 19.3 ± 2.7 viral genomic copies/μL when using RNA extracted samples obtained from nasopharyngeal swabs collected in guanidine-containing viral transport medium. Typical RT-LAMP reactions were performed at 65 ºC for 30 min. When compared to RT-qPCR, up to Ct value 32, RT-LAMP presented 97% (87.4-99.4% 95% CI) sensitivity and 100% (86.2-100%) specificity for SARS-CoV-2 RNA detection targeting N gene. No cross-reactivity was detected when testing other non-SARS-CoV virus, confirming high specificity. The test is compatible with primary RNA extraction free samples. We also demonstrated that colorimetric RT-LAMP can detect SARS-CoV-2 variants of concern (VOC) and variants of interest (VOI), such as variants occurring in Brazil named P.1, P.2, B.1.1.374 and B.1.1.371. The method meets point-of-care requirements and can be deployed in the field for high-throughput COVID-19 testing campaigns, especially in countries where COVID-19 testing efforts are far from ideal to tackle the pandemics. Although RT-qPCR is considered the gold standard for SARS-CoV-2 RNA detection, it requires expensive equipments, infrastructure and highly trained personnel. In contrast, RT-LAMP emerges as an affordable, inexpensive and simple alternative for SARS-CoV-2 molecular detection that can be applied to massive COVID-19 testing campaigns and save lives.

## INTRODUCTION

Emerging viral infections continue to pose a major threat to global public health. In the past decades different viral emergencies have been reported including the severe acute respiratory syndrome coronavirus (SARS-CoV), H1N1 influenza, Middle East respiratory syndrome coronavirus (MERS-CoV), Ebola vírus, Zika virus and most recently the new coronavirus has been described, which cause COVID-19 (1,2). COVID-19 has as etiologic agent the <u>s</u>evere <u>a</u>cute <u>r</u>espiratory <u>s</u>yndrome <u>co</u>rona<u>v</u>irus 2 (SARS-CoV-2), which belongs to the *Coronaviridae* family, *Betacoronavirus* genus (3,4). People with COVID-19 have a wide range of symptoms reported such as fever, cough, anosmia, ageusia, headache, fatigue, muscle or body aches, sore throat, shortness of breath or difficulty breathing. Some of these symptoms help spread the virus, however human-to-human transmission from infected individuals with no or mild symptoms has been extensively reported (5,6). This outbreak has spread rapidly, as of May 2021, there were over 165 million confirmed COVID-19 cases with over 3,4 million deaths recorded worldwide (https://coronavirus.jhu.edu/). Isolation and quarantine of infected individuals is essential to viral spread and community dissemination of airborne pathogens and requiring an accurate, fast, affordable, readily available tests for massive population testing. In contrast do antibody detection, which may take weeks after the onset of the infection. Detection of viral RNA is the best way to confirm the acute infection phase, the most important phase for viral shedding, so that rationally managed social distancing and lockdown can be implemented (1,7).

Quantitative reverse transcription-polymerase chain reaction (RT-qPCR) is considered the gold standard method for SARS-CoV-2 RNA detection, mainly targeting combinations of viral genome regions that codes for nucleocapsid protein (N), envelope protein (E), RNA-dependent RNA polymerase (RdRp) and other targets on the open reading frame (ORF1ab) (8). Although RT-qPCR assays have played an important role un the SARS-CoV-2 diagnosis; the technique has limitations for massive population testing such as processing time, the requirement of sophisticated equipment, infrastructure and highly trained personnel, as well as costly reagents with high demand and shortages around the world. Thus, developing complementary, inexpensive point-of-care (PoC) methods that are rapid, simple, allowing the use of alternative reagents for COVID-19 diagnosis test, is urgently needed. Methods gathering these features can make affordable massive testing campaigns, including contact tracing strategies in highly dense countries, saving lives (9– 17). In this regard reverse transcription loop-mediated isothermal amplification (RT-LAMP) has been shown to be an affordable technique applied to detect different pathogens (18,19). RT-LAMP has been used during Ebola outbreak (20,21) and for tracking Zika virus (22) or Wolbachia (23) in Brazilian mosquitoes. The method relies on specific DNA amplification at constant temperature without the need for sophisticated thermal cyclers (24). The amplified products can be visually detected by magnesium pyrophosphate precipitation; fluorescence emission by DNA intercalating dyes; agarose gel electrophoresis; lateral flow immunochromatography; magnesium chelating color indicators and pH-dependent colorimetric reaction that changes from fuchsia (pink) to yellow (positive result) due to proton release during nucleic acid amplification (25) (Figure 1). The possibility of accessing results by naked eye, made RT-LAMP an exciting alternative that facilitates the use of COVID-19 molecular testing. Simple, scalable, cost-effective RT-LAMP-based alternatives for SARS-CoV-2 detection, has emerged during pandemics including protocols for viral inactivation, quick run, RNA extraction-free and LAMP-associated CRISPR/Cas strategies (10,11,14,16,17,26–32). On April 14^th^, 2020, the RT-LAMP received the emergency use authorization from the United States Food and Drug Administration for SARS-CoV-2 detection in COVID-19 diagnostics.

**Figure 1.**
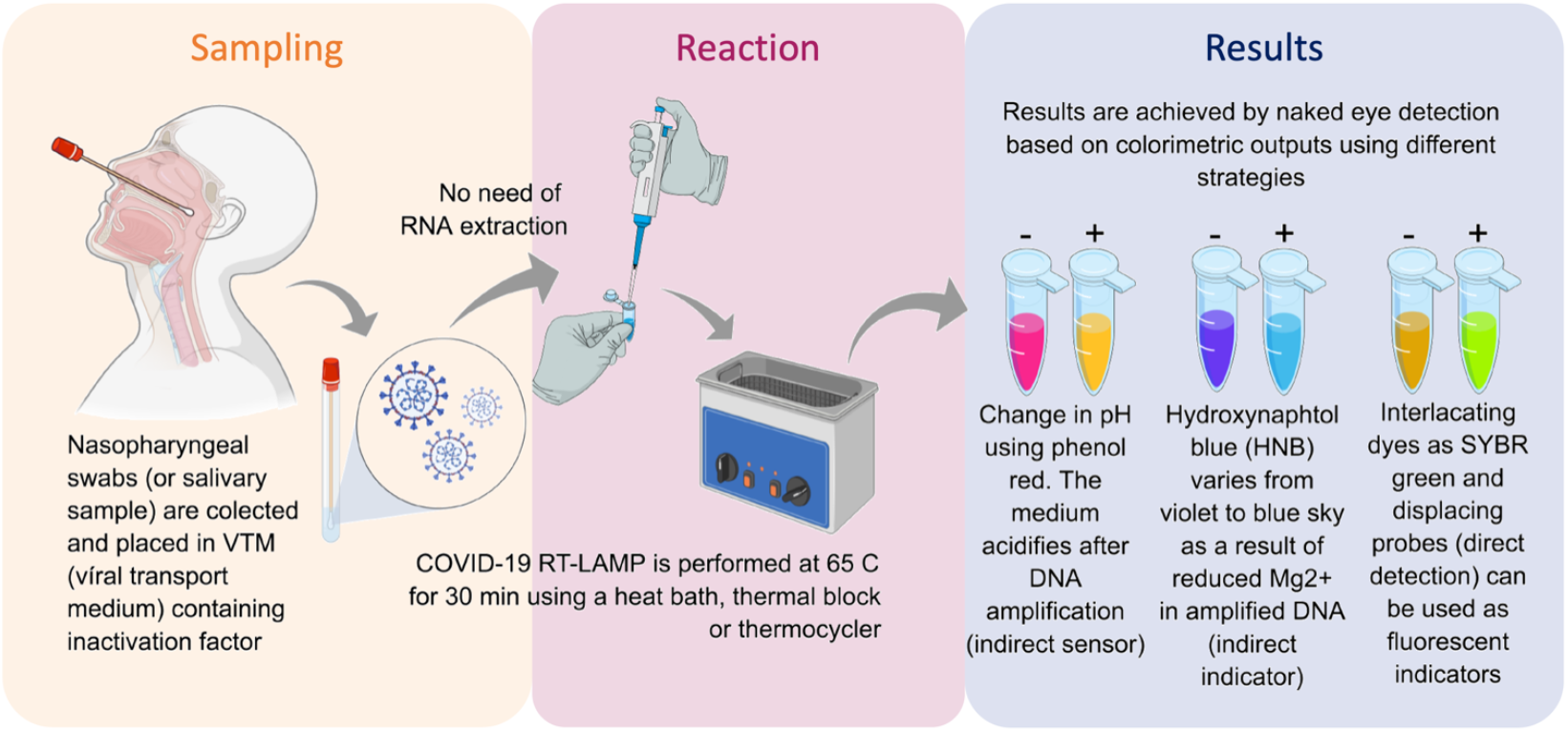
Reverse transcription loop-mediated isothermal amplification (RT-LAMP) for SARS-CoV-2 RNA detection and COVID-19 testing. Inactivated saliva samples or from nasopharyngeal swabs can processed for RNA extraction previously or be directly added to RT-LAMP reaction. Colorimetric output can be achieved by different sensors and can be read by naked eye. The whole procedure is rapid, simple and do not requires complex infrastructures.

In this study, we validate a colorimetric RT-LAMP assay to detect SARS-CoV-2 RNA in clinical samples collected in different parts of Brazil, including samples with known SARS-CoV-2 variants of interest and concern. After testing primers used by RT-LAMP SARS-CoV-2 RNA detection targeting different regions, best results were achieved when using *N* gene or *N/E* genes-based strategies. One hundred nasopharyngeal swabs collected in a guanidine-containing viral transport medium (VTM) (33) from symptomatic hospitalized patients were tested. The clinical validation revealed a sensitivity of 97% (87.4-99.4 95% CI) with samples ranging Ct values from 15 to 32 with 100% specificity. The use of RNA extraction-free samples was also tested, although there is a loss in sensitivity. We also demonstrated that RT-LAMP is affordable for the detection of more transmissible SARS-CoV-2 variants encompassing a number of genomic nucleotide changes. Part of the results presented here are the research basis of OmniLAMP^®^ SARS-CoV-2 kit which was approved by the Brazilian Heath Regulatory Agency for COVID-19 molecular testing (Anvisa nº: 10009010368) as an alternative for massive decentralized diagnostic in Brazil, that records the third-highest COVID-19 cases number worldwide (https://coronavirus.jhu.edu). Together with vaccination, RT-LAMP for COVID-19 diagnosis could help to improve better life quality during pandemics, offering an alternative molecular testing for monitoring lock-down measures; traveling restrictions; the return of universities, schools, kindergartens; sport leagues (including the presence of audience, in the Olympics for example) activities with worldwide impact.

## RESULTS

### RT-LAMP targeting SARS-CoV-2 N and E genes can detect as low as 19 viral copies/µL

In order to access absolute analytical sensitivity of the colorimetric RT-LAMP for SARS-CoV-2 detection, we calculated the limit of detection (LoD), which is the lowest detectable concentration of viral nucleic acid – here represented in viral copies per microliter (cps/µL), which was determined based on a calibration curve from a known copy number load standard E gene-harboring plasmid. Purified SARS-CoV-2, obtained from infected Vero E6 cells revealed a LoD equivalent to 0.44 ± 0.2 cps/µL while RNA obtained from clinical samples (nasopharyngeal swab in viral transport medium – VTM) resulted in a LoD of 19.3 ± 2.7 cps/µL. Validation was performed using clinical samples, confirming the LoD by colorimetric RT-LAMP as well as by the visualization of the amplified DNA in agarose gel (Figure 2).

**Figure 2.**
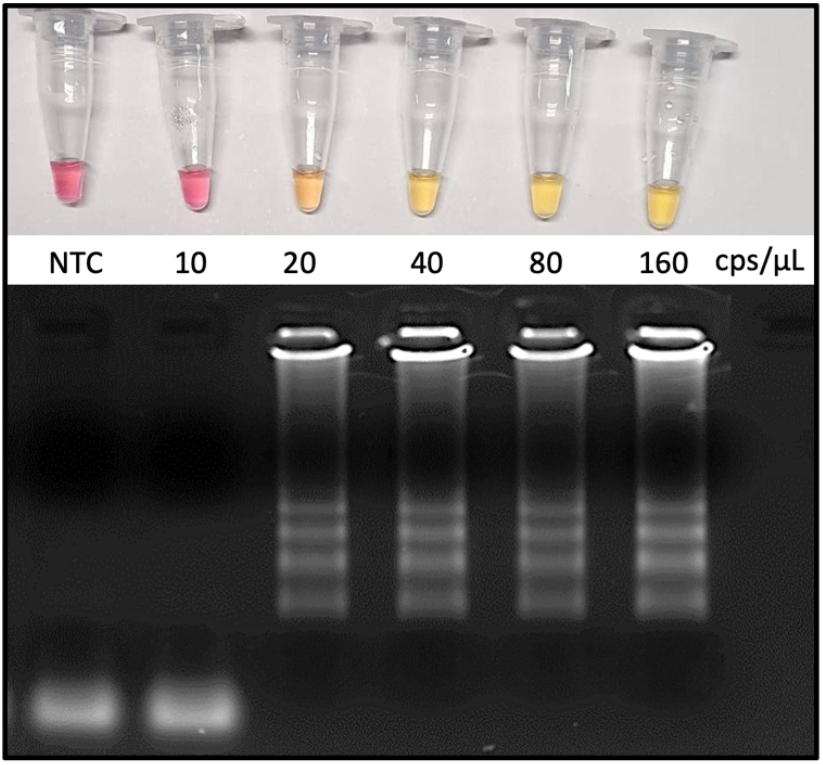
Analytical sensitivity as revealed by the limit of detection (LoD). RNA was extracted from VTM-nasopharyngeal swab and the genome viral copies input was calculated based on SARS-CoV-2 E gene-harboring plasmid (Bioclin #K228-1) calibration curve. RT-LAMP reaction was performed at 65 °C during 30 min using WarmStart^®^ colorimetric master LAMP mix (NEB #M1800) in 20 μL final volume (upper panel). Amplicons were resolved in 2% agarose gel and stained with GelRed^®^ (Biotium #41003) to confirm DNA amplification (bottom panel). cps/µL: viral genome copies per microliter; NTC: non-template control; VTM: viral transport medium (Bioclin #G092-1).

### SARS-CoV-2 detection by RT-LAMP on clinical samples having N gene as target present 100% specificity while sensitivity varies from 100-87% depending on the viral load

The diagnostic accuracy for RT-LAMP was compared to the “gold standard” technique RT-qPCR. The relative sensitivity was accessed in a panel of 100 clinical specimens from nasopharyngeal swab collected in VTM, including 60 positive and 40 negative samples according to the colorimetric RT-LAMP output (Figure 3) that were previously characterized by RT-qPCR (Table 1). The colorimetric output was correlated with the visualization of amplified DNA after agarose gel electrophoresis.

**Table 1.**
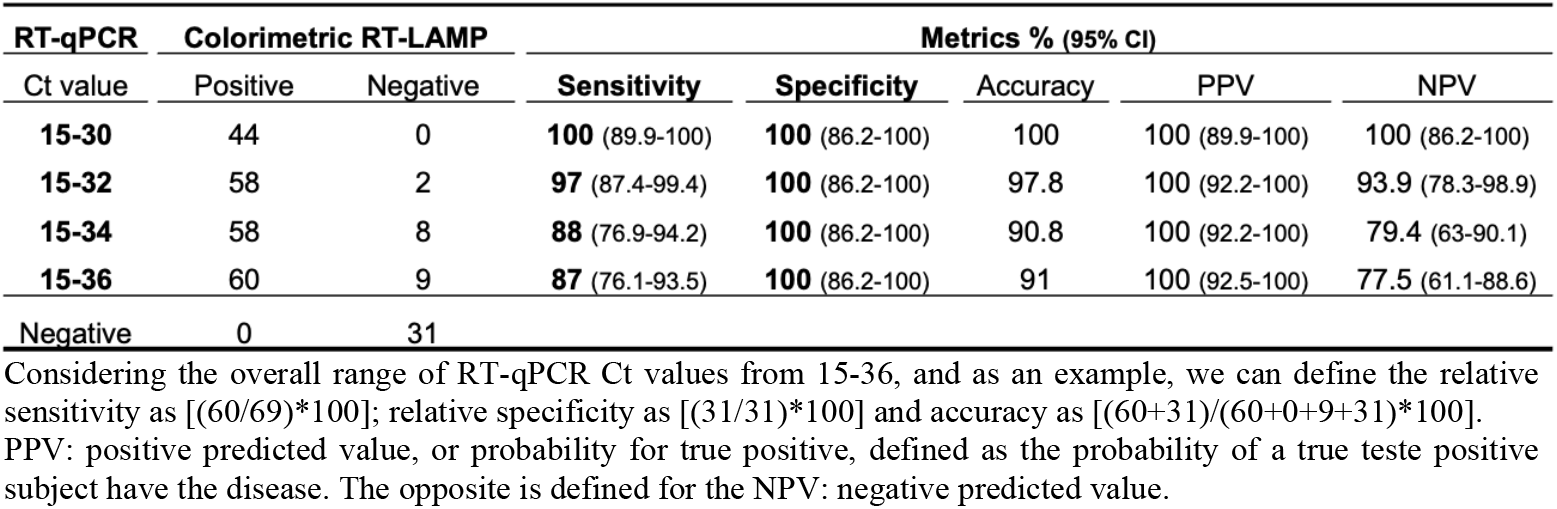
Comparison of clinimetric parameters between colorimetric RT-LAMP and RT-qPCR on the detection of SARS-CoV-2 for molecular diagnosis of COVID-19

| RT-qPCR | Colorimetric RT-LAMP |  | Metrics % (95% CI) |  |  |  |  |
| --- | --- | --- | --- | --- | --- | --- | --- |
| Ct value | Positive | Negative | Sensitivity | Specificity | Accuracy | PPV | NPV |
| 15-30 | 44 | 0 | 100 (89.9-100) | 100 (86.2-100) | 100 | 100 (89.9-100) | 100 (86.2-100) |
| 15-32 | 58 | 2 | 97 (87.4-99.4) | 100 (86.2-100) | 97.8 | 100 (92.2-100) | 93.9 (78.3-98.9) |
| 15-34 | 58 | 8 | 88 (76.9-94.2) | 100 (86.2-100) | 90.8 | 100 (92.2-100) | 79.4 (63-90.1) |
| 15-36 | 60 | 9 | 87 (76.1-93.5) | 100 (86.2-100) | 91 | 100 (92.5-100) | 77.5 (61.1-88.6) |
| Negative | 0 | 31 |  |  |  |  |  |

Considering the overall range of RT-qPCR Ct values from 15-36, and as an example, we can define the relative sensitivity as $[(60/69)*100]$ ; relative specificity as $[(31/31)*100]$ and accuracy as $[(60+31)/(60+0+9+31)*100]$ . PPV: positive predicted value, or probability for true positive, defined as the probability of a true teste positive subject have the disease. The opposite is defined for the NPV: negative predicted value.

**Figure 3.**
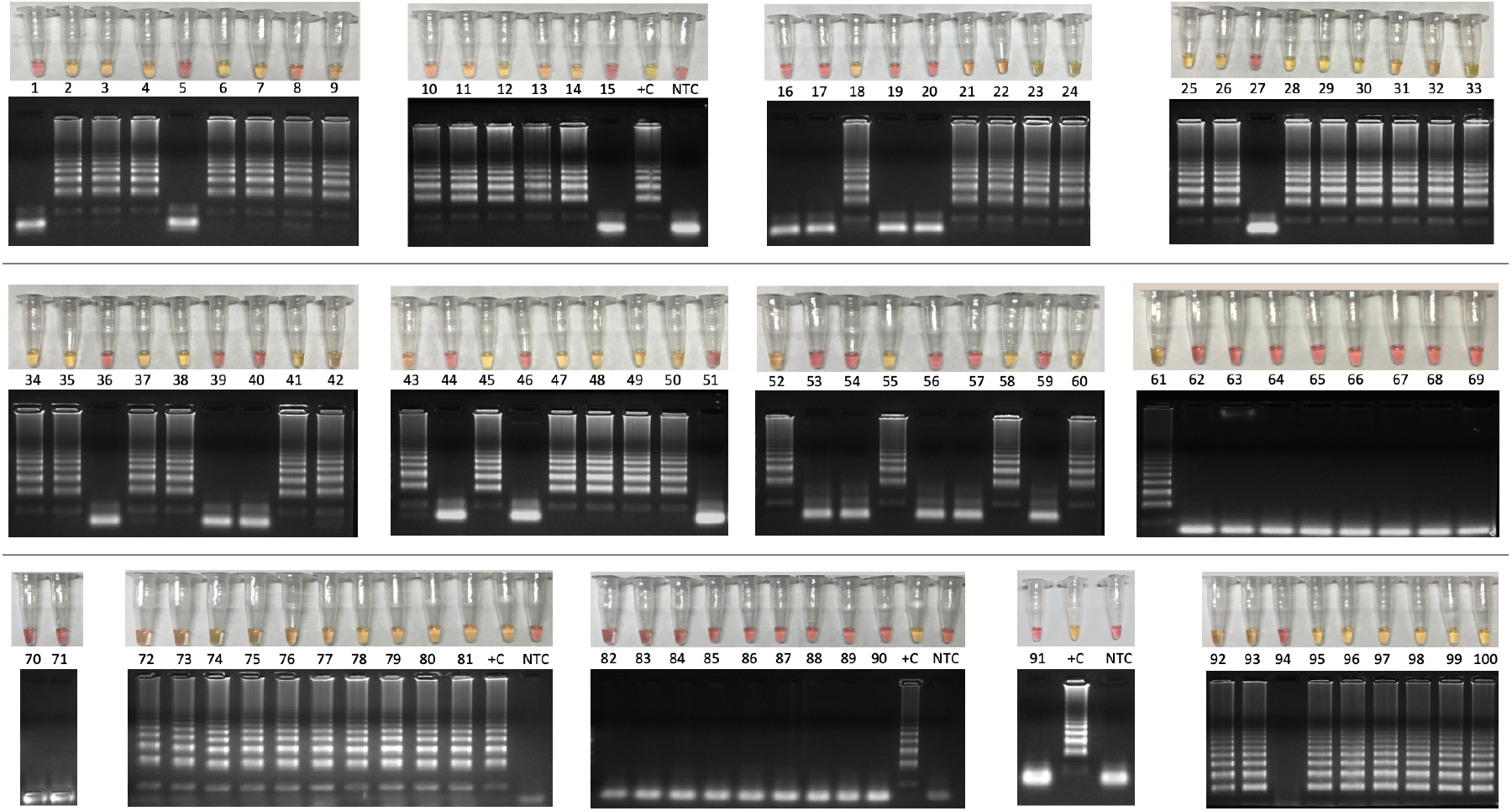
Colorimetric RT-LAMP for COVID-19 diagnosis validation using one hundred clinical samples. Clinical samples were collected from symptomatic patients by nasopharyngeal swabs in partnership with CT-Vacinas/UFMG, Belo Horizonte, Brazil. Samples were obtained from different parts including Brazilian Southeast and Northeast regions. The reaction was performed at 65 °C during 30 min using WarmStart^®^ colorimetric LAMP master mix (NEB #M1800) in 20 μL final volume. The RT-LAMP reaction targeted SARS-CoV-2 N gene. Yellow content indicate positive reaction while pink pattern reveal non-reagent samples. Amplicons were resolved in 2% agarose gel and stained with GelRed^®^ (Biotium #41003) to confirm DNA amplification. Latter pattern confirmed specific SARS-CoV-2 amplification that matches with yellow output tubes which is not observed in pink non-reagent tests. +C: positive control using RNA extracted from laboratory-Vero E6 cultured inactivated SARS-CoV-2. NTC: non-template control.

The overall accuracy of colorimetric RT-LAMP compared to RT-qPCR was 91%, considering RT-qPCR Ct values ranging from 15 to 36, with relative sensitivity of 87% (95% CI 76.1-93.5%) and 100% (95% CI 86.2-100%) specificity (Table 1). However, considering samples with equivalent RT-qPCR Ct value ≤ 32, RT-LAMP sensitivity is 97% (95% CI 87.4-99.4%) and reaches 100% (95% CI 90-100%) in samples with Ct value ?? 30, while specificity is always 100% (Table 1), which means there are no false positive hits. It is noteworthy that some samples with RT-qPCR Ct > 32 had false negatives results for RT-LAMP (Table 1 and Figure 4A). However, other four samples were detected as positive on RT-LAMP with RT-qPCR Ct values ranging from 32-36 (Figure 4A). Analysis by receiver operating characteristic (ROC) curve confirmed high sensitivity and specificity at RT-PCR equivalent Ct value 31.8 for RT-LAMP on COVID-19 diagnostics (Figure 4B)

**Figure 4.**
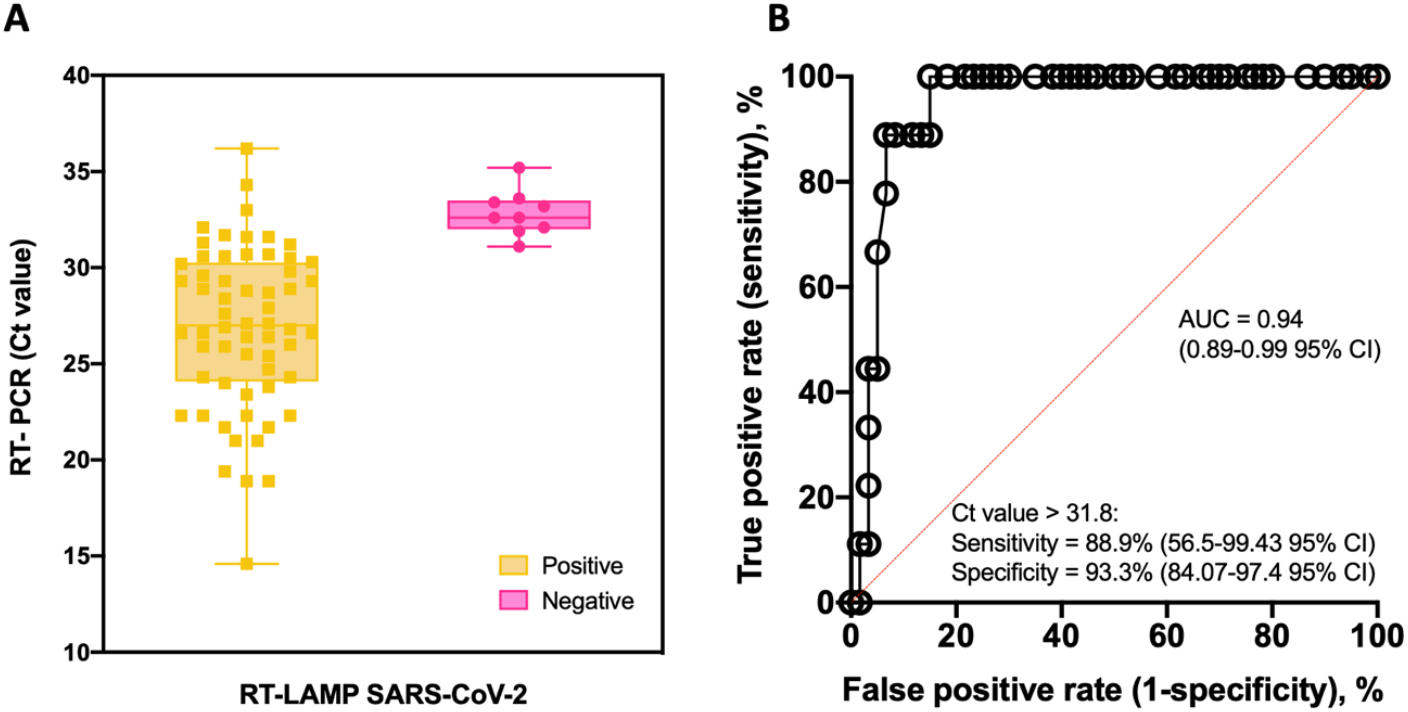
Colorimetric RT-LAMP for SARS-CoV-2 RNA detection. A) Box and whisker representation of colorimetric RT-LAMP SARS-CoV-2 positive and negative output (x axis) plotted in function of their respective RT-PCR Ct values (y axis). Nine false negative samples were detected on RT-LAMP after Ct 32 despite other four being positive from Cts ranging from 32-36. B) Receiver operating characteristic (ROC) curve constructed based on data presented in A. As summarized in Table 1, high sensitivity and specificity values were obtained at the predicted cut-off.

### RT-LAMP targeting SARS-CoV-2 N gene do not cross-reacted with other viruses, including respiratory ones

The analytical specificity was confirmed by performing RT-LAMP for SARS-CoV-2 on putative cross-reacting viruses such as pathogens that colonizes the human upper respiratory tract or that are associated with seasonal outbreaks in Brazil. None of the tested viruses (human Influenza A virus/H1N1; Influenza B virus; human respiratory syncytial virus, Dengue, Zika and Chikungunya viruses) presented cross-reactivity on RT-LAMP using N gene as SARS-CoV-2 target (Figure 5). It reinforces the high specificity observed on clinical validation with no false positive results (Figure 3). Thermodynamic and alignment analyses were performed on SARS-CoV-2 N, E and RdRp RT-LAMP primer sets, revealing that there is no cross-reactivity over more than 300 non-SARS coronaviruses-derived genomes (Table S1).

**Figure 5.**
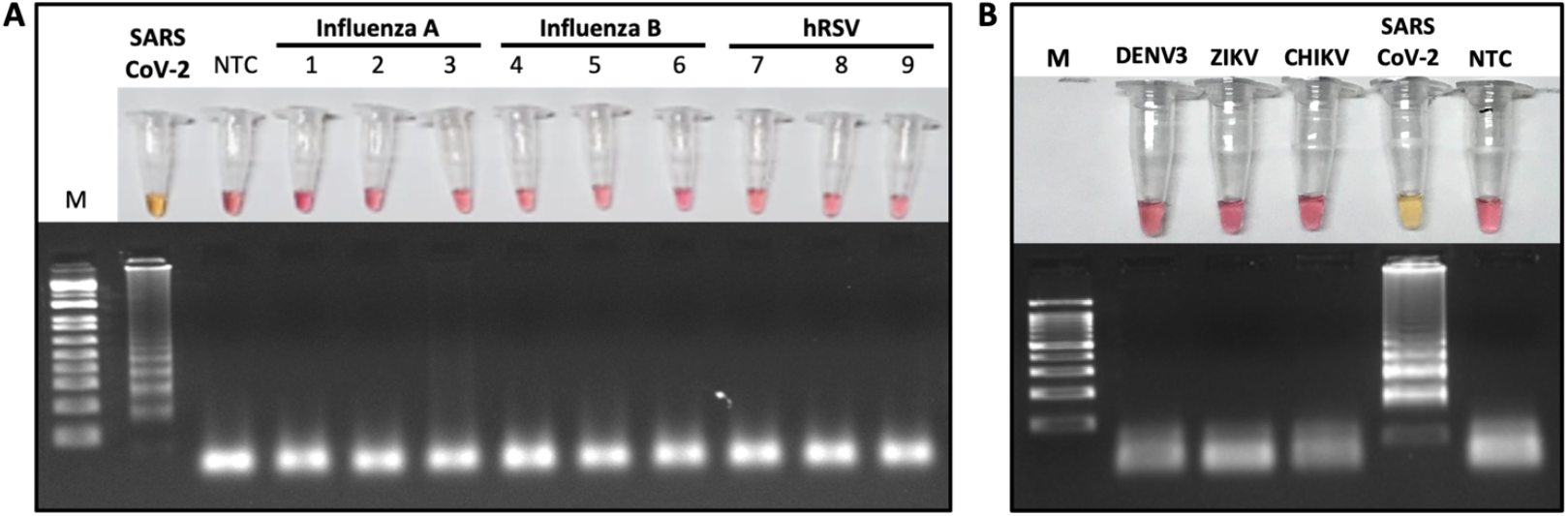
Microbial cross-reactivity assay to test SARS-CoV-2 RT-LAMP analytical sensitivity. The test was performed using potentially cross-reacting respiratory viruses (A) or local occurring arboviruses (B). RT-LAMP reaction was performed at 65 °C during 30 min, with additional 10 min, to confirm the absence of cross-reactivity when using SARS-CoV-2 N gene as target. The assay was performed using the WarmStart^®^ colorimetric LAMP 2x master mix (NEB #M1800). Yellow (positive) reaction is only observed when the template is SARS-CoV-2 viral RNA. hRSV: human respiratory syncytial virus; NTC: non-template control; M: molecular size marker. RT-LAMP amplification products were resolved in 2% agarose gel and stained with GelRed^®^ (Biotium #41003) to confirm DNA amplification. DENV3: Dengue virus serotype 3; ZIKV: Zika virus; CHIKV: Chikungunya virus; Influenza A (H1N1/H3N2); Influenza B (Yamagata/Victoria).

### SARS-CoV-2 N gene favors sensitivity when compared to E and RdRp genes as target for singleplex RT-LAMP

Six clinical samples previously confirmed as SARS-CoV-2 positive by RT-qPCR were subclassified as presenting low, medium or high Ct values for E gene as target. All of them were tested by colorimetric RT-LAMP in independent reactions to test the performance of N, E and RdRp genes as target to detect SARS-CoV-2. The low Ct values (18.9 and 21.7) samples were positive for all tested primer sets, while E and RdRp genes started to present false negative results from medium (26.6 and 28.4) Ct values (Figure 6). It indicates that the SARS-CoV-2 N gene is a better target for colorimetric RT-LAMP, detecting viral RNA in samples with equivalent RT-qPCR Ct values over 30 (Figure 6).

**Figure 6.**
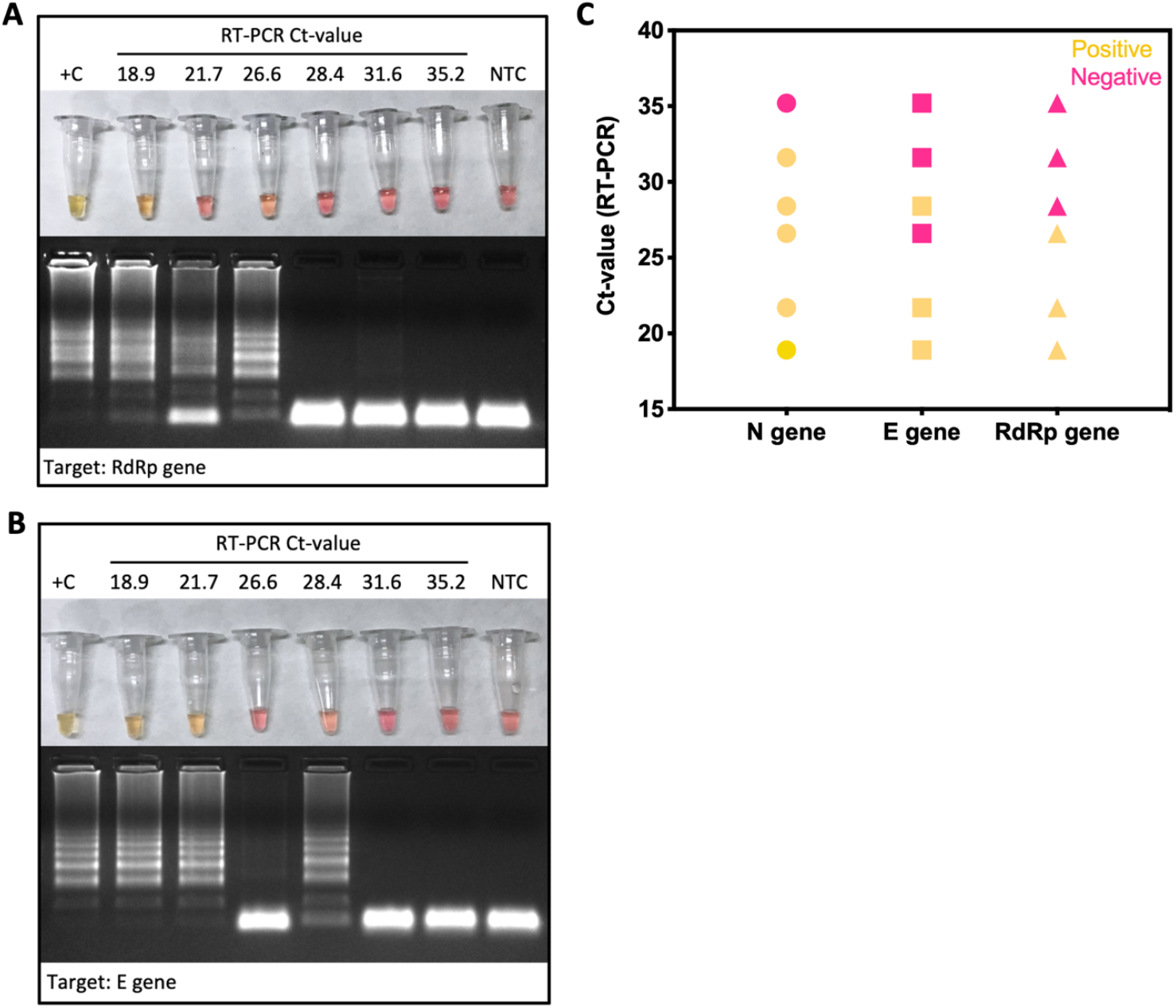
Colorimetric RT-LAMP for SARS-CoV-2 detection using genes N, E and RdRp as target. Selected SARS-CoV-2 positive clinical samples by RT-qPCR were classified as low (Ct 18.9 and 21.7); medium (Ct 26.6 and 28.4) and high (Ct 31.6 and 35.2) Ct values for E gene. They were included as input for colorimetric RT-LAMP reaction using primers targeting N, RdRp (A) and E genes (B). RT-LAMP SARS-CoV-2 false negative samples are more frequent when using E and RdRp genes as target (C). RT-LAMP reaction was performed at 65 °C during 30 min, using the WarmStart^®^ colorimetric LAMP 2x master mix (NEB #M1800). RT-LAMP amplification products were resolved in 2% agarose gel and stained with GelRed^®^ (Biotium #41003) to confirm DNA amplification. +C: positive control using SARS-CoV-2 RNA extracted from laboratory-cultured inactivated SARS-CoV-2. NTC: non-template control.

Colorimetric RT-LAMP sensitivity depends on the set of LAMP primers that can vary even within the same target. When RT-LAMP was performed on low viral load samples (Ct value for E gene ranging from 31.8 – 36.2), the N gene_Set1 was able to identify 4 out of 12 (33.3%) true positive samples. In contrast, N gene_Set2 or primer multiplex strategy (N gene Set1/Set2) allowed the detection of 11/12 (91,6%) true positive samples (Supplemental Table S2).

### Colorimetric RT-LAMP can be performed on clinical samples without RNA extraction

RT-LAMP performed in clinical samples, without any chemical or physical pre-treatment or RNA extraction, returned positive with three out of five positive samples (Figure 7A). In this assay we used laboratory-cultured and inactivated SARS-CoV-2 and clinical samples without previous RNA extraction, showing that it is possible to use direct patients’ samples without pre-processing (Figure 7A). However, this should be taken with caution, as crude clinical samples may contain interferents. Previous heat-inactivation can be used to reduce this possibility. Here, only 1 μL of 1:10 solution of hydrochloride guanidine-containing VTM from nasopharyngeal swabs was added as a template to the SARS-CoV-2 LAMP reaction. Further analyses are being performed to establish the method sensitivity and feasibility for massive patient screening. All five samples had had previous RNA extraction, for RT-PCR analysis, supporting that extraction process can increase detection sensitivity. We also tested the incubation time at 65 °C reaction temperature. All SARS-CoV-2 control samples turned reaction color from fuchsia to yellow as indicative of DNA amplification, confirming positive reaction from the earliest time point tested (Figure 7B). In all tested intervals non-template controls were pink/fuchsia (negative) as expected, without any spurious late amplification, as confirmed by agarose gel electrophoresis showing no amplification bands on it (Fig 7B).

**Figure 7.**
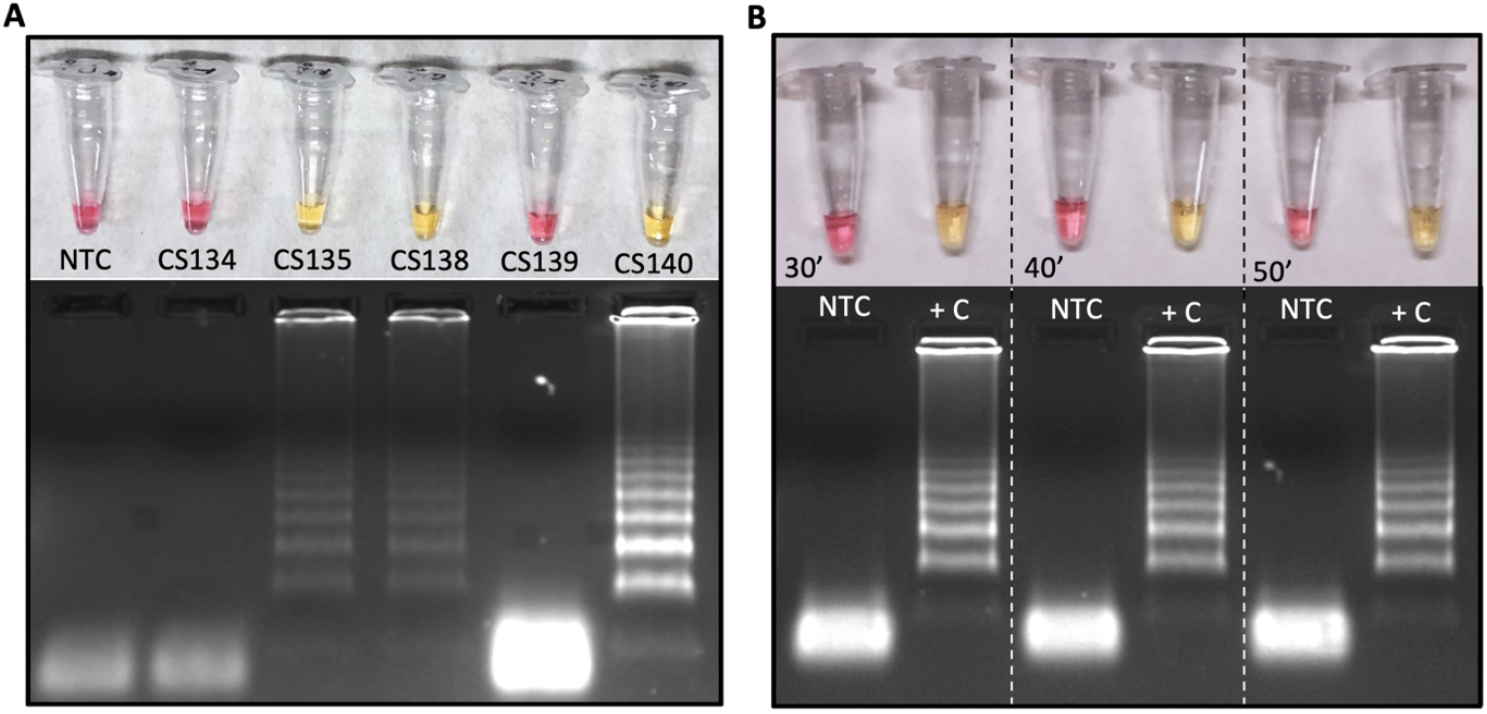
Colorimetric RT-LAMP to detect SAR-CoV-2 in RNA extraction-free clinical samples (A) or laboratory-cultured virus (B). Clinical samples were derived from nasopharyngeal swabs placed on guanidine-containing viral transport medium, diluted 1:10. The RT-PCR Ct values for SARS-CoV-2 based on E gene are: CS134 = 31.8; CS135 = 15.3; CS138 = 18.4; CS139 = 21.7 and CS140 = 24.6. RT-LAMP reaction was performed in 20 μL final volume, incubated at 65 °C during 30, 40 or 50 min (inactivated virus) using WarmStart^®^ colorimetric LAMP master mix (NEB #M1800). Both, clinical samples or viruses, are RNA extraction-free samples. The amplification products (amplicons) were migrated in agarose gel at 2% to confirm amplification, as indicated by the characteristic ladder highlighted by GelRed^®^ staining. NTC: non-template control; CS: clinical sample; +C: positive control.

### Colorimetric RT-LAMP allows the detection of new SARS-CoV-2 variants of interest (VOI) and concern (VOC)

As a worldwide concern, SARS-CoV-2 VOI and VOCs molecular detection could fail when applying S region-based RT-qPCR diagnostic methods due to mutations that would prevent primer annealing. In order to provide experimental evidences that RT-LAMP as a powerful molecular tool for detecting SARS-CoV-2 RNA, including VOC and VOI, we performed the tests on clinical samples that were previously identified as VOC/VOI by complete genome sequencing. All tested variants, including P.1 (B.1.1.28.1) and P.2 (B.1.1.28.2) – originally reported in Brazil – were detected in colorimetric SARS-CoV-2 RT-LAMP (Figure 8), either by N gene alone as target or by multiplex strategy using N2/E1 primer set, indicating that none of the mutant polymorphisms prevents specific primer annealing on RT-LAMP COVID-19 diagnosis (Figure 8).

**Figure 8.**
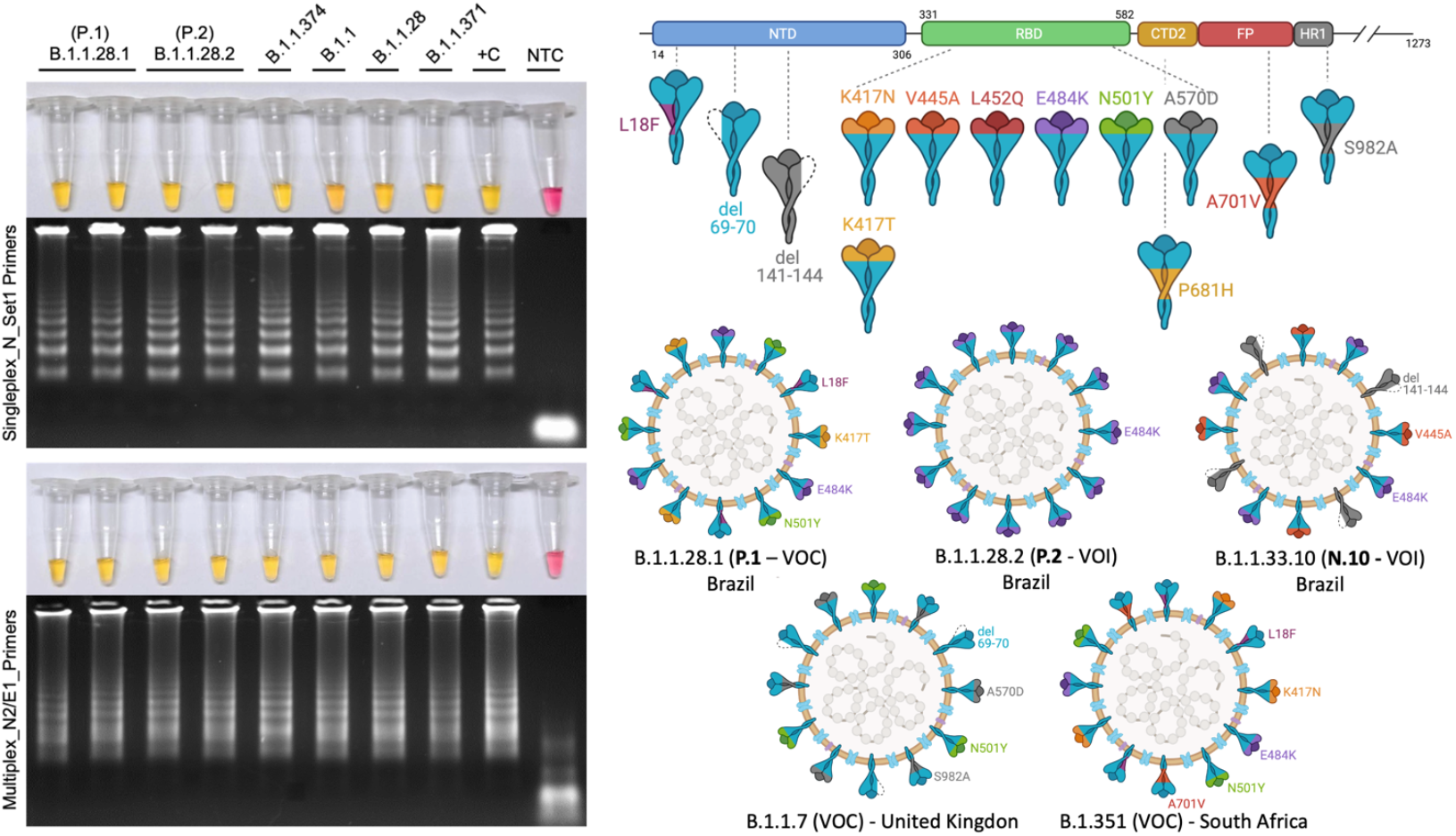
Colorimetric RT-LAMP allows the detection of SARS-CoV-2 variants of concern (VOC) and interest (VOI). RT-LAMP reaction was performed at 65 °C for 30 min, using the WarmStart^®^ colorimetric LAMP 2x master mix (NEB #M1804), using either SARS-CoV-2 N gene set1 primers (upper left panel) or multiplex N2/E1 primer sets (down left panel). The amplicons were migrated in agarose gel at 2% to confirm amplification, as indicated by the characteristic ladder highlighted by GelRed^®^ staining. NTC: non-template control; CS: clinical sample; +C: positive control. The right hand panel shows a schematic representation of SARS-CoV-2 spike protein (upper) and highlights as representative SARS-CoV-2 virions, the main marker mutations present in Brazilian VOC P.1, VOI P.2 and N.10 as well as the VOC B.1.1.7 and B1.3.51 firstly reported in the United Kingdom and South Africa, respectively. K417N: lysine to asparagine substitution at position 417 of spike protein at the receptor biding domain (RBD); V445A: valine to alanine substitution at position 445 and so on. L: Leucine; Q: glutamine; E: glutamic acid; Y: tyrosine; T: threonine; P: proline; H: histidine; D: aspartic acid; S: serine; F: phenylalanine. del: deletion. Segments of SARS-CoV-2 protein NTD: N-terminal domain; CTD2: C-terminal domain 2 or C terminus of S1 fragment after furin cleavage; FP: fusion peptide; HR1: heptad repeat region 1. SARS-CoV-2 variants were previously sequenced. Variants of interest B.1.1.371 and B.1.1.374 were first reported in Saudi Arabia and Finland, respectively (https://cov-lineages.org/).

## DISCUSSION

The COVID-19 pandemics demanded a rapid global response in massive diagnostic solution to face the worldwide crisis. In this context, the RT-qPCR – considered the gold-standard technique for SARS-CoV-2 RNA detection – requires high-cost equipments, trained personnel and specialized laboratory structure. In addition, during COVID-19 pandemics, several health care centers and private laboratories competed for RT-qPCR kits and related products to meet the high diagnostic demand. In order to overcome the lacking of molecular testing and provide affordable alternatives, RT-LAMP had become one of the main hopes. Due to its simplicity, accuracy comparable with RT-qPCR to detect SARS-CoV-2 RNA, does not require PCR machine and naked eye readable colorimetric output, RT-LAMP was the focus of massive testing campaigns (14,16,28). This screening strategy is compatible with: home, primary care clinics, point of entry (borders), schools, universities, sport leagues, companies and can help to achieve a safe back to work and quarantine monitoring (10,14,16,28,30). Since April 14^th^, 2020 the U.S. Food and Drugs Administration (FDA) issued the emergence use authorization of Color SARS-CoV-2 RT-LAMP Diagnostic Assay from Color Health, Inc. (EUA number: EUA200539).

In order to provide an affordable SARS-CoV-2 detection tools, we validate a colorimetric RT-LAMP for the COVID-19 diagnosis using clinical samples collected from different parts of Brazil. The country has a flawed screening performance, testing less than 220 individuals per 1,000 people (May 2021) (https://www.worldometers.info/coronavirus) where the majority of tests rely on antibodies-based rapid tests which are not the most reliable and recommended for mass screening and decision making to control local outbreaks. The test sensitivity of RT-LAMP, which is comparable to gold standard RT-qPCR and clearly relies on: the target choice, incubation time, viral load (asymptomatic patients, days of symptoms, correct sampling), output reading, sample integrity and quality (viral transport media, sample storage condition, pre-analytical treatments, extraction procedure, crude RNA extraction free samples) and sample type (nasal, nasopharyngeal, saliva, sputum, gargle lavage) (Table 2).

**Table 2.**
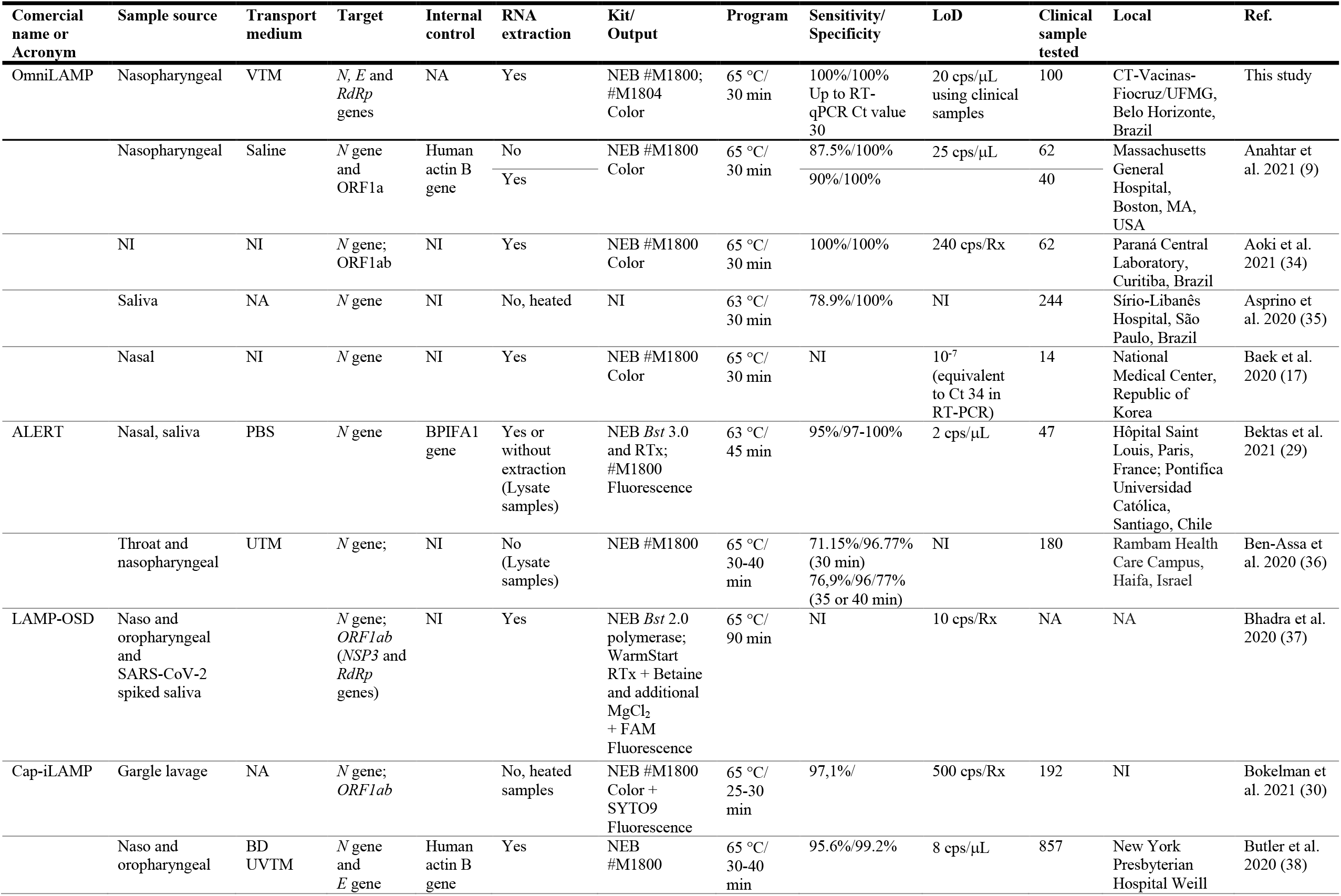
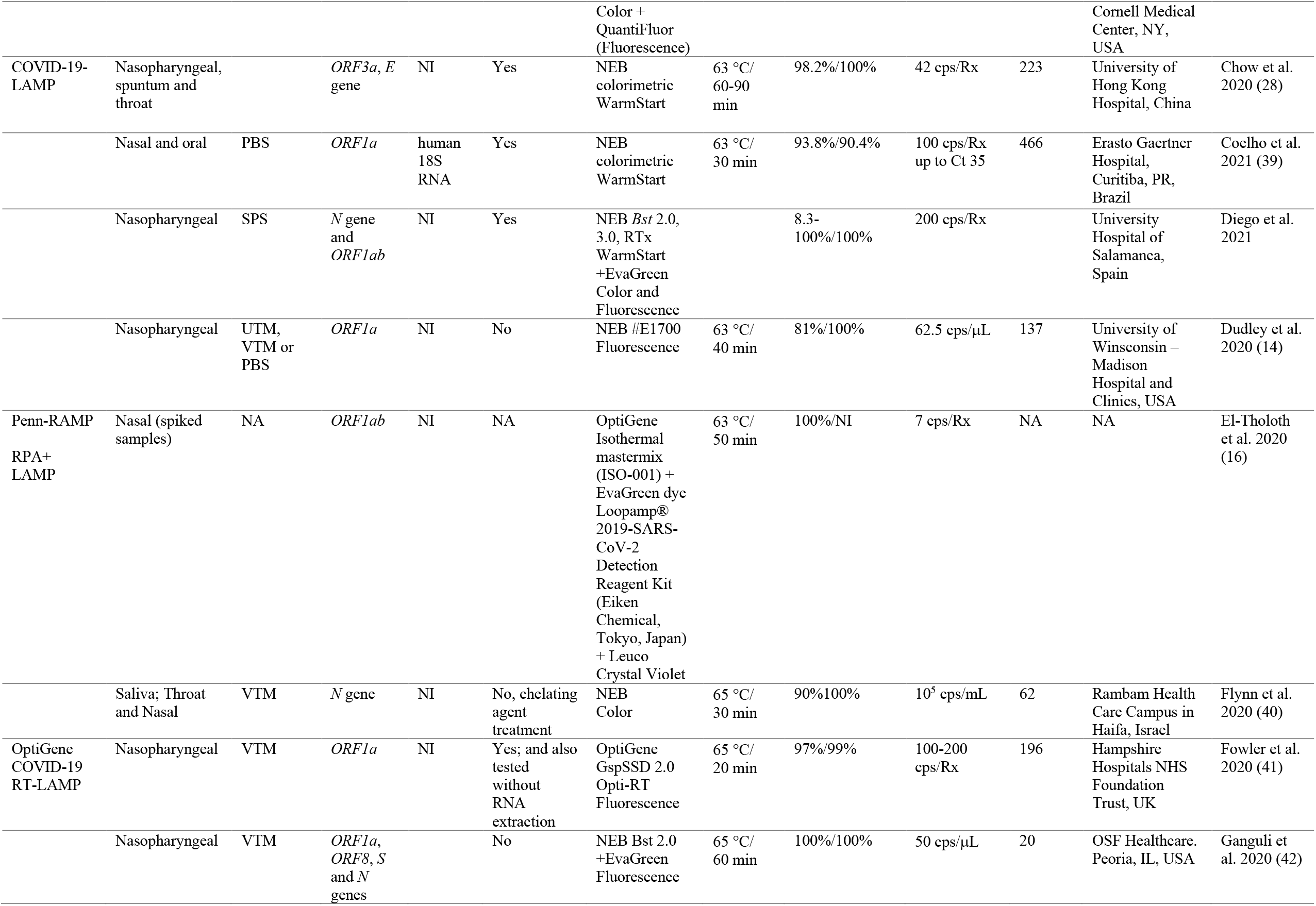
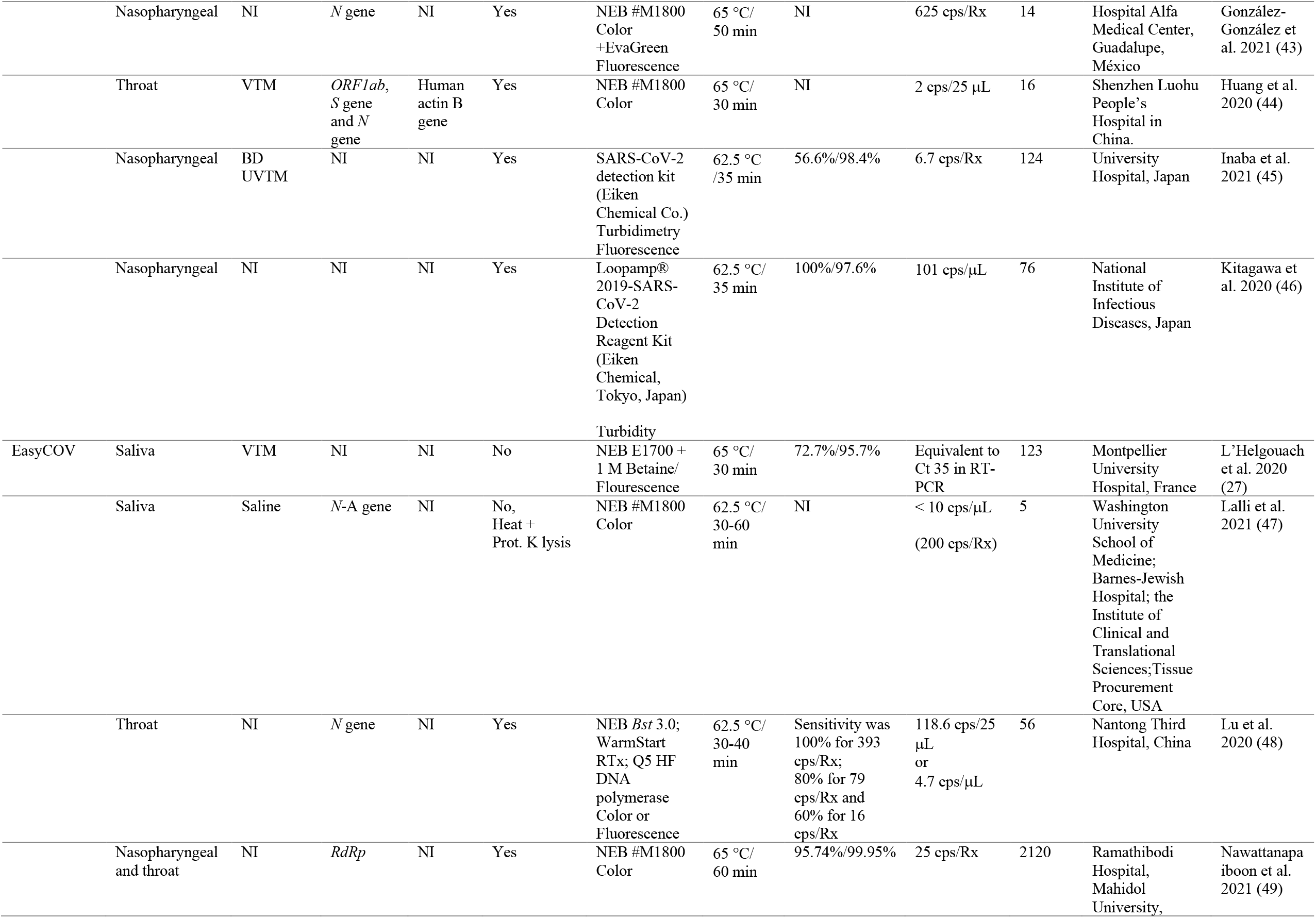
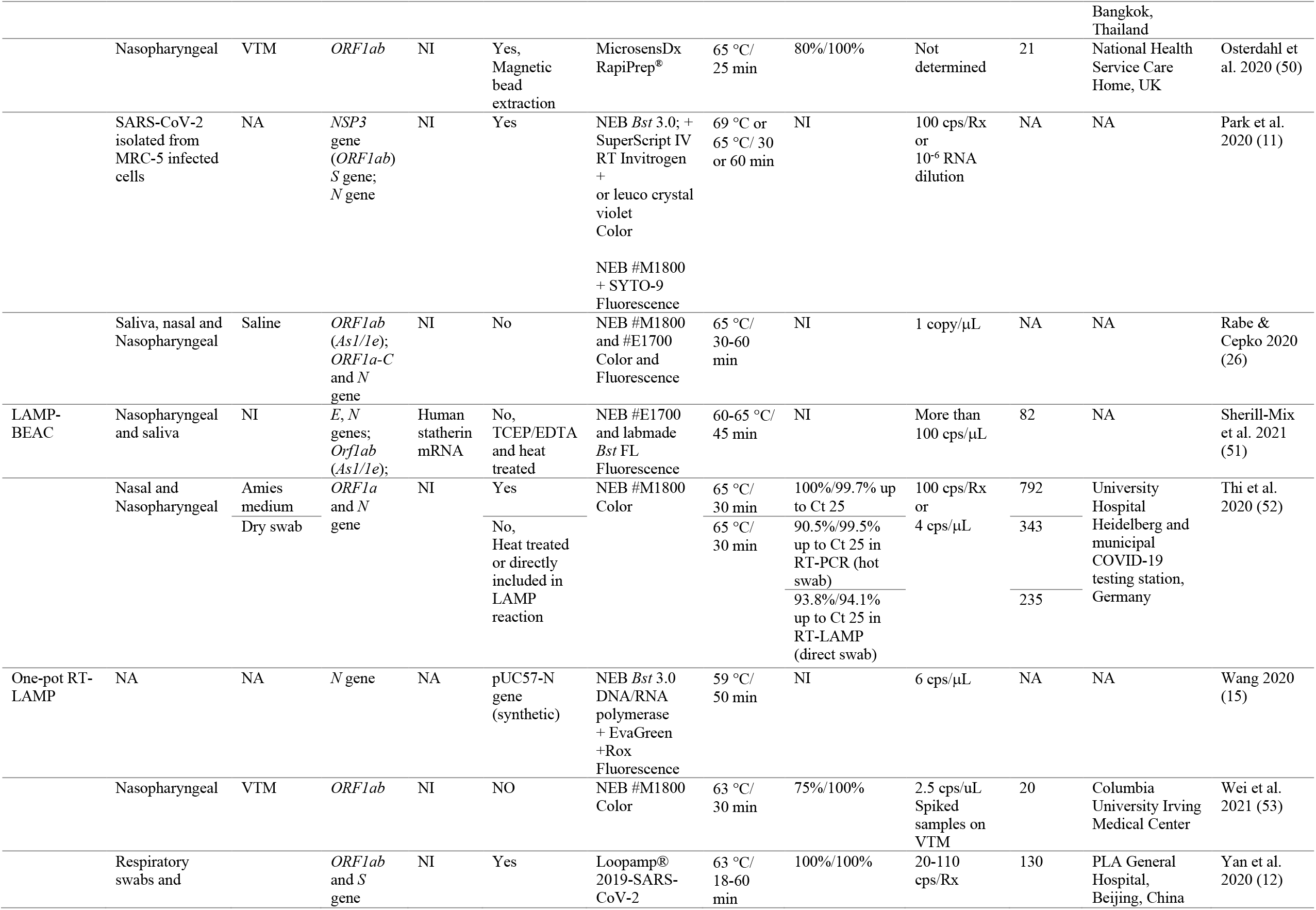
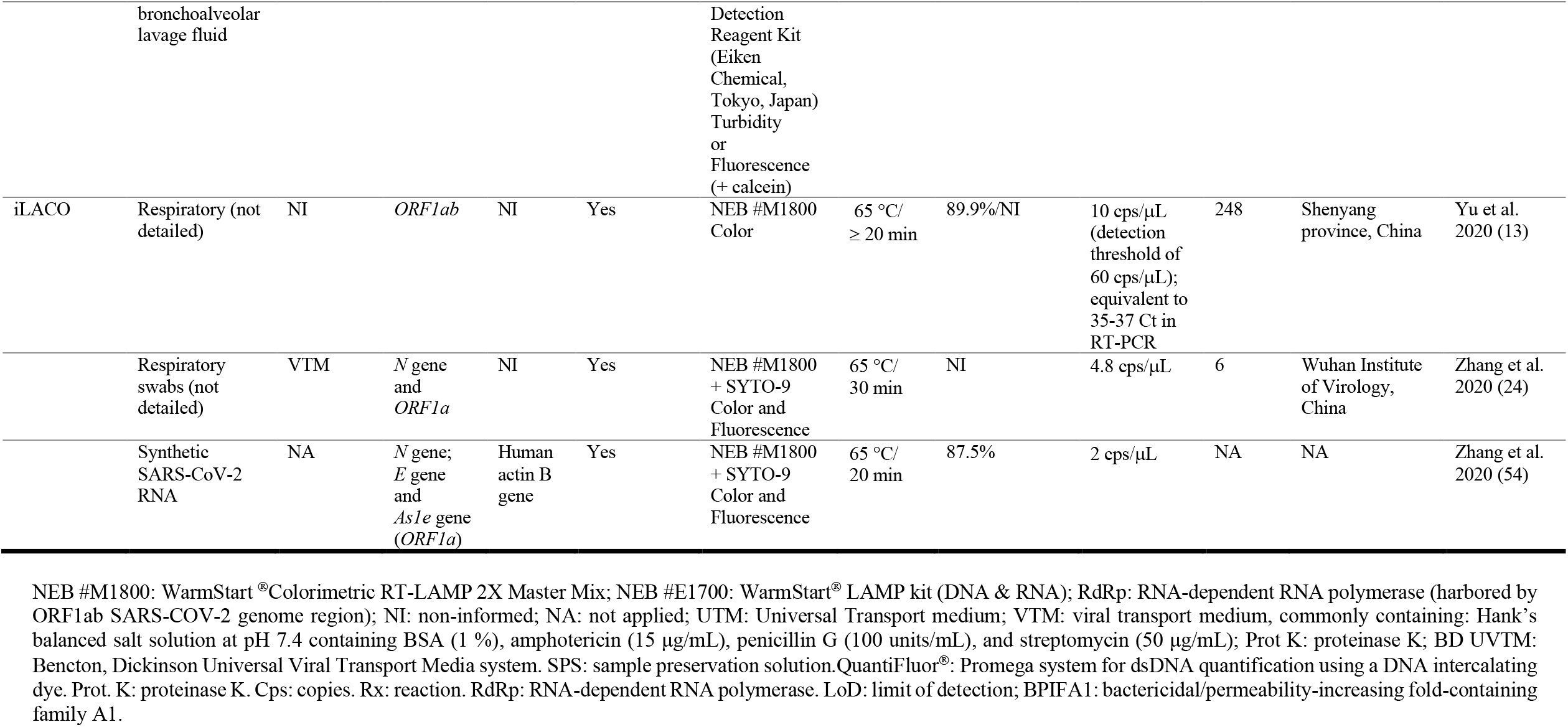
Comparison of SARS-CoV-2 RT-LAMP solutions, including key parameters on clinical validation.

Upon RNA extraction from nasopharyngeal swab-derived clinical samples we found a LoD of 20 viral genomic copies/µL, confirming previous studies based on N SARS-CoV-2 target (9,30,43). It is worth noting that when using non-clinical SARS-CoV-2 extracted RNA or synthetic target, the LoD reaches less than 0.5 cps/µL. This can be explained by the presence of interferents such as, VTM, host cells and enzymes that could reduce the yield (14,49). In this regard, one has to be careful when interpreting LoD calculated using non-clinical samples. Nevertheless, extracted samples are rich enough in viral genomic copies to meet clinically relevant levels of SARS-CoV-2.

Clinical validation of RT-LAMP for COVID-19 diagnosis relies on calculating parameters such as sensitivity, specificity, PPV, NPV and accuracy compared to the gold standard RT-qPCR. We have to be careful when associating the RT-LAMP sensitivity and indirect assumption on RT-qPCR viral load is not straightforward due to some technical concerns. It is well accepted that Ct values can be representative of viral load. However, this parameter could lead to misinterpretation when comparing different kits, targets and non-standardized samples. A survey conducted by the College of American Pathologists on more than 700 laboratories, reported a variation as much as 14 cycles among different methods on the same batch material. Single laboratories using different platforms and targets in SARS-CoV-2 molecular testing can represent a potential variability on Ct-values (55). Considering previous convergent reports and presuming different targets and platforms, the data from the literature shows that with a RT-qPCR Ct 30 cut-off where RT-LAMP sensitivity for SARS-CoV-2 detection is close to 100% (Ben-Assa et al., 2020; Lamb et al., 2020; L’Helgouach et al., 2020; Smyrlaki et al., 2020; Lalli et al., 2021; Newman et al., 2021; Yang et al., 2021)(39,52,56,57) and eventually with a higher threshold Ct 35 as well (27,39). Indeed, we confirm that up to Ct 30 RT-LAMP returned 100% sensitivity for SARS-CoV-2 detection, reaching 97% and 88% when considering Ct values up to 32 and 34 respectively. Curiously, Kim and cols. (2021) found that samples from hospitalized patients presenting Ct value of 28.4 or less were infective to cell human culture (58), an evidence – based on *in vitro* extrapolation – that RT-LAMP sensitivity is compatible with threshold of infectivity. This reinforces that simple, robust and reliable RT-LAMP meets clinical requirements, presenting similar COVID-19 diagnostic accuracy as RT-qPCR (45,50,59).

The choice of SARS-CoV-2 genomic target plays an important role when selecting the RT-LAMP method for COVID-19 diagnosis. Several research groups have tested different regions on SARS-CoV-2 genome with the potential to generate RT-LAMP primers. Once, the majority of primers were designed using the open source software Primer Explorer, it is expected that at some point, the default algorithm returned the same result or overlapping regions, independently identified in a context where molecular biology scientists everywhere in the world are working to tackle COVID-19 (Figure 9). According to our data compilation, *N* gene and *ORF1ab* regions (overlapping *NSP3, As1e* and *RdRp*-coding sequences) were the most frequent targets chosen for SARS-CoV-2 RT-LAMP (Table 2 and Figure 9). Ganguli and cols. (2020) and Zhang and cols. (2020) arrived at the same conclusion when selecting the SARS-CoV-2 *N* gene-targeting primer set after confirming better performances for RNA viral detection when compared to other targets (42,54). When testing *N, E* and *RdRp* genes in true positive – previously RT-qPCR characterized clinical samples – we observed more false negative outputs from assays using *E* and *RdRp* genes, corroborating what was previously reported. We also highlight that primer subsets within the same N target gene, can contribute differentially to RT-LAMP test sensitivity (Supplementary Table S2). Furthermore, multiplexing different primer sets is encouraged in order to increase sensitivity (Figure 9) (54,60,61).

**Figure 9.**
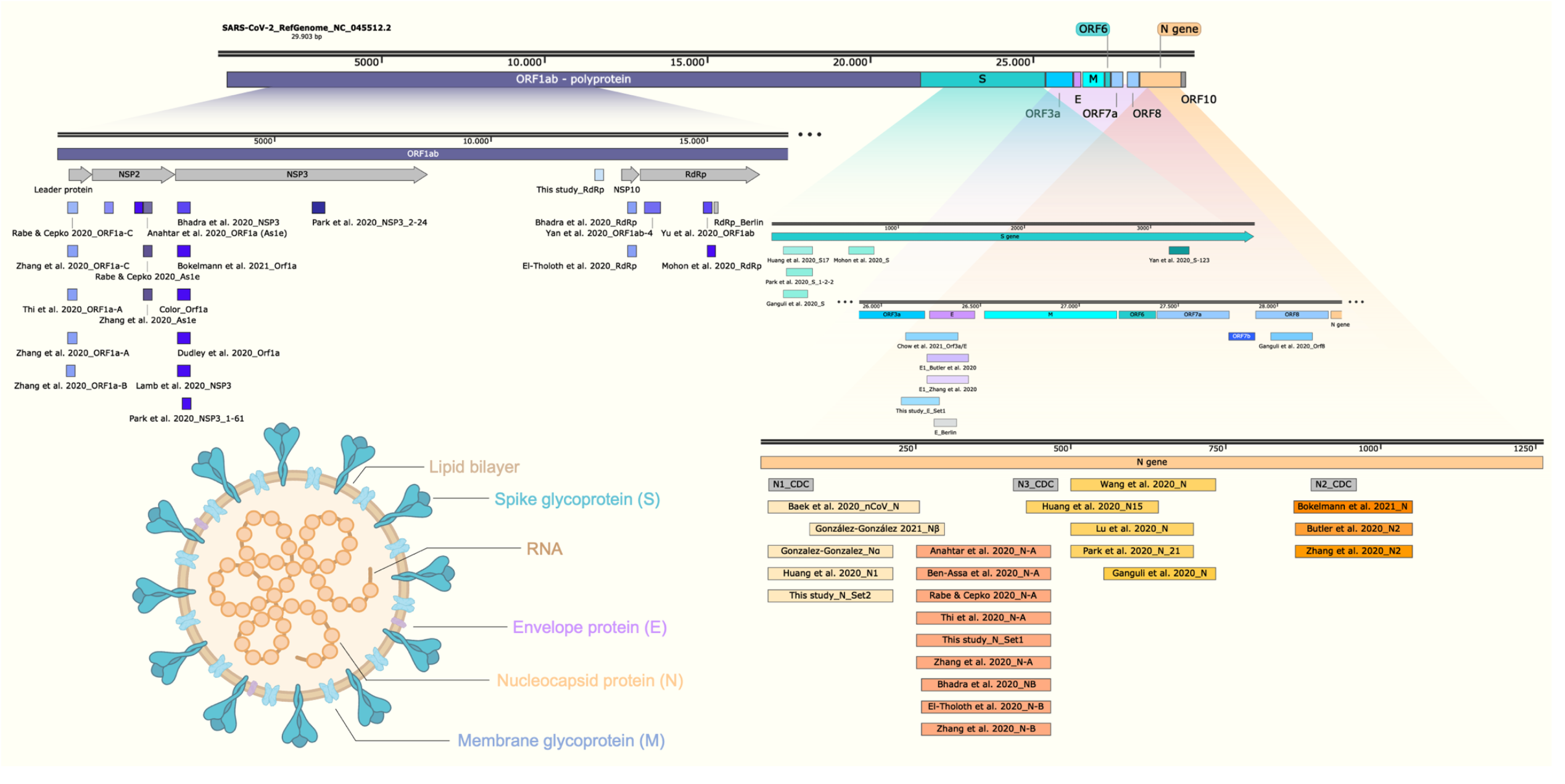
Schematic representation of SARS-CoV-2 genome indicating the amplicons for the COVID-19 molecular diagnostics by RT-LAMP. Structural representation of SARS-CoV-2 virion shows the main particle parts. LAMP primer regions are indicated as previously reported (9,12– 14,16,17,24,26,28,30,36–38,42,43,52,63,65,85,86); ORF: open reading frame; RdRp: RNA-dependent RNA polymerase; NSP: non-structural protein. Schematic representation created using Snap Gene Viewer software version 5.0.7; N1, N2 and N3_CDC correspond to the amplicons for SARS-CoV-2 detection by RT-PCR (86).

Another important (almost) neglected point, is the fact that, although inspired by RT-qPCR target selection, few SARS-CoV-2 RT-LAMP approaches reported an internal control target to confirm the presence of human RNA and monitor sampling or extraction process (39). Wilson-Davies and colleagues (2021) pointed out that the lack of amplification can happen for different reasons concerning the whole reaction, a specific well or due to inhibitory substances; highlighting the importance of including internal control even before nucleic acid extraction, in order to be considered a reliable SARS-CoV-2 LAMP assay (62). We have not include internal controls in the clinical validation presented here, since all accessed samples had been previously characterized by RT-qPCR, including human RNAse P as housekeeping gene. However, in the current OmniLAMP^®^ assay, we included human β-actin RNA (rACTB) as internal control. Other constitutive targets for SARS-CoV-2 RT-LAMP include BPIFA1 (29), human 18S RNA (39) and Statherin RNA (51) (Table 2).

Similar to its high sensitivity, obtained in our and by other studies, the SARS-CoV-2 RT-LAMP specificity is undoubtebly high and is frequently reported as 100% without any cross-reactivity with other respiratory or SARS-CoV unrelated viruses (11,28,34,49,63). We also confirm that the SARS-CoV-2 RT-LAMP solution presented here is highly specific and does not cross-react with Brazilian occurring seasonal Influenza A and B, hRSV or arboviruses.

Despite the advantages presented by purified and nucleic acid enriched samples for SARS-CoV-2 RT-LAMP, RNA extraction-free protocols have attracted attention since they can be non-invasive saliva-based; do not require additional steps and equipment and fulfill point-of-sampling requirements. Indeed, the pre-analytical phase on RT-LAMP is the bottleneck for point-of-care applications. For this reason, several studies highlighted the feasibility of primary RNA extraction-free approaches for SARS-CoV-2 RNA detection (8,9,14,27,35,36,47,53,57,64). Pre-treatment of saliva samples includes heat sample inactivation and the use of lysis/stabilizing buffers that can contain proteinase K, TCEP, EDTA, DTT could help the viral RNA assessment maintaining its integrity (27,36,47,65–68). Caution must be taken when running colorimetric RT-LAMP since pre-treatment could interfere on result outputs. One of the main limitations for direct sample test by colorimetric RT-LAMP based on pH-sensing is the false positive results upon input sample addition (previous to amplification), due to naturally acidic samples (30,52). To prevent spurious amplification due to the presence of DNA from oral microbiome, food or host cells on primary samples, Bokelmann and cols (2021) treated samples with λ exonuclease that acts by preferentially digesting 5’-phosphorylated DNA leaving non-phosphorylated primers or LAMP products intact (30). Here we shown preliminary results on RNA extraction-free, pre-treatment free, primary 10x diluted hydrochloride guanidine-containing VTM nasopharyngeal samples directly accessed to compared colorimetric results. Three out of five RT-qPCR true positives directly accessed samples returned positive yellow output on colorimetric RT-LAMP for SARS-CoV-2 detection. This provides clues on the use of unextracted samples for massive COVID-19 testing campaigns with a trade-off on cost-benefits for limit of detection and test sensitivity. Most high and medium viral load samples will be detected on unextracted protocols. However, to meet RT-qPCR detection sensitivity levels, this requires some type of purification step and RNA concentration (11,24,31).

We are currently observing rapid converging evolution of SARS-CoV-2 during COVID-19 pandemics worldwide. Several reports alert for the emergence of variants of concern and interests such the B.1.1.7, first detected in England (ECDC – threat assessment brief on December 20^th^ 2020) (69); B.1.351 initially reported from South Africa (70); B.1.1.28.1 (P.1) that was identified in Japan but obtained from a traveler from Brazil (33) and more recently the B.1.617.1 and B.1.617.2 VOC detected in India, but that already spread to South America (Argentina’s Health Ministry official report). The regional selection of SARS-CoV-2 VOC is associated with higher transmissibility, mortality and reduced neutralizing antibody response (71–74). In Brazil, we observed the emergence of different SARS-CoV-2 VOC and VOI, including P.1, B.1.28.2 (P.2) (75,76); B.1.1.33.9 (N.9) (77); B.1.1.33.10 (N.10) (78,79). A plethora of mutations is observed in these variants, including N501Y, E484K/Q, K417N/T, A570D and the Δ69-70 at the SARS-CoV-2 S protein sequence, which was associated with detection failures by S-target RT-qPCR methods (80). For SARS-CoV-2 RT-LAMP detection, few studies selected S-coding protein region as a target (Figure 9). In addition, isothermal amplification for SARS-CoV-2 RNA detection strategies is commonly addressed as multiplex targeted, making RT-LAMP a good choice even for SARS-CoV-2 variants detection. Indeed, here we reported that singleplex *N* gene-based or multiplex *N2*/*E1*-based RT-LAMP was able to perfectly detect VOC and VOI circulating in Brazil such as P.1, P.2, B.1.1.374 and B.1.1.371 (Figure 8), the two latter firstly detected in Finland and Saudi Arabia (https://cov-lineages.org). Recent efforts made by Sherrill-Mix and cols (2021) showed a beacon-based RT-LAMP strategy designed to precisely identify B.1.1.7 SARS-CoV-2 variant (51,81), a promising tool not only for massive screening but also to monitor VOC/VOI SARS-CoV-2 spreading.

## CONCLUSIONS

The colorimetric RT-LAMP is a reliable molecular tool for detecting SARS-CoV-2, providing rapid and easy to read results; compatible with high-throughput screenings and point-of-care requirements. This test is especially important for nations with a poor diagnostic conditions, like in Brazil, where RT-qPCR COVID-19 diagnostic is far from ideal to control disease spreading. The RT-LAMP sensitivity can be equivalent to those reported from the gold standard RT-qPCR method and also present 100% specificity. Results are commonly obtained after 30 min reaction and if needed, additional 20 min was not associated with spurious unspecific amplification. Alternative directly accessed samples from diluted guanidine-containing VTM (nasopharyngeal) without any pre-treatment interventions can also be used, however, with lower sensitivity. Sample collection in guanidine-containing VTM has been described as a useful strategy to avoid contamination of health care workers during sample manipulation. RT-LAMP primer selection can directly interfere on sensitivity being N gene the best target for SARS-CoV-2 RNA detection with less false negative results, especially in low viral load samples. Colorimetric RT-LAMP is also compatible with detecting SARS-CoV-2 variants of interest and concern, being robust to cope with the monitoring of emerging new SARS-CoV-2 variants and that can be easily adapted. We thus, reinforce and recommend the use of RT-LAMP for massive testing as a decentralized point-of-care alternative for SARS-CoV-2 containment and to tackle COVID-19.

## MATERIAL AND METHODS

### Clinical samples, RT-qPCR and ethical statement

One hundred clinical SARS-CoV-2 samples were obtained from symptomatic hospitalized patients from different parts of Brazil. Nasopharyngeal swabs were collected and maintained in 2 mL VTM (Bioclin, Belo Horizonte, Brazil #G092-1) at room temperature until RNA extraction or direct dilution for LAMP reaction. The VTM contains guanidine chloride as inactivation agent and to preserve viral RNA. All procedures were performed inside a biosafety level 2 cabinet. RNA extraction was performed using the QIAamp^®^ Viral RNA Mini Kit (Qiagen #52906), following manufacturer instructions. The molecular diagnostic routine was performed by RT-qPCR using the SARS-CoV-2 commercial kits produced at Fundação Oswaldo Cruz (Kit Molecular SARS-CoV-2 E/RP, from Bio-Manguinhos/Fiocruz – based on Charité/Berlin protocol and Kit Biomol OneStep/COVID-19 from IBMP/Fiocruz – based on China/CDC protocol with recommended targets polyprotein ORF1ab and N gene). RT-qPCR was carried out using the 7500, ViiA 7 real-time PCR systems (Applied Biosystems, Foster City, CA. USA) or the dual-channel Open qPCR machine (Chai, Santa Clara, CA, USA), following the temperature program profile of 95 °C for 3 min, followed by 40 cycles of amplification (95 °C/15 s and 60 °C/1 min). Influenza and hRSV samples were kindly provided by IOM/FUNED and the arboviruses samples are part of the collection from the laboratório de Imunologia de Doenças Virais at Oswaldo Cruz Foundation. All procedures involving human participants, collection and use of clinical samples and data were in accordance with ethical standards and approved by the human research ethics committee under license protocol number: 4051614; CAAE (certificate of presentation for ethical appreciation): 31984720300005091. SARS-CoV-2 variants of concern and variants of interest included in this study were isolated from symptomatic patients (Ct value < 25, using E gene as target on RT-qPCR – Kit Molecular SARS-CoV-2 E/RP Bio-Manguinhos Fiocruz), in the State Pernambuco, Northeast Brazil (82).

### RT-LAMP primer design

RT-LAMP primers were designed based on SARS-CoV-2 reference genome (GenBank accession NC_045512.2) using the open source software Primer Explorer V5 (https://primerexplorer.jp/lampv5e) or the NEB LAMP primer design tool (https://lamp.neb.com). The free energy (??G) of selected primers was less than -4 kcal/mol, as a parameter chosen based on oligo stability (83). The set of primers used in this study are listed in Table 1 and additional information can be found in Figure 9 and Supplementary Figures S3, S4 and S5. We designed and validated different LAMP primer sets, such as N gene Set1 and Set2 that appeared in other independent researches (Figure 9 and Table 3). N2 and E1 primer sets were previously designed by Zhang et al. (2020) (54). The oligos were purchased from Integrated DNA technologies (IDT, Coralville, IA, USA) and from Exxtend (Paulínia, SP, Brazil). All oligos were synthesized at 25 nanomole scale and purified by standard desalting. Thermodynamic evaluation of primers targeting SARS-CoV-2 N, E and RdRp genes were performed as previously described (84). Briefly, hybridization temperature of F3, FIP (F1c+F2), BIP (B1c + B2), LF and LB primer sets were calculated upon aligning to SARS-CoV or other coronavirus (non-SARS) genomes, considering potential mismatches. The SARS-CoV-2 coverage for each primer was also obtained (Supplementary Table S1).

**Table 3.**
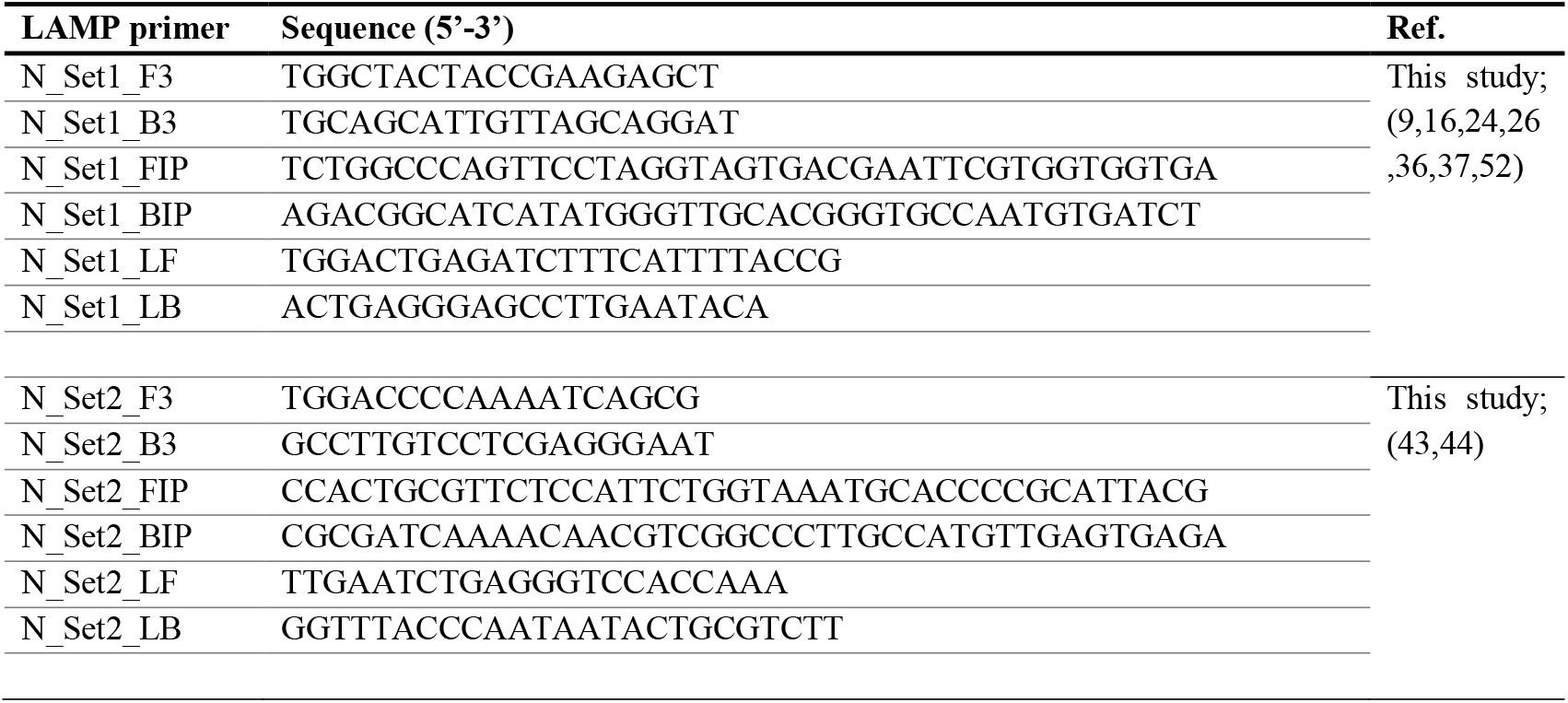
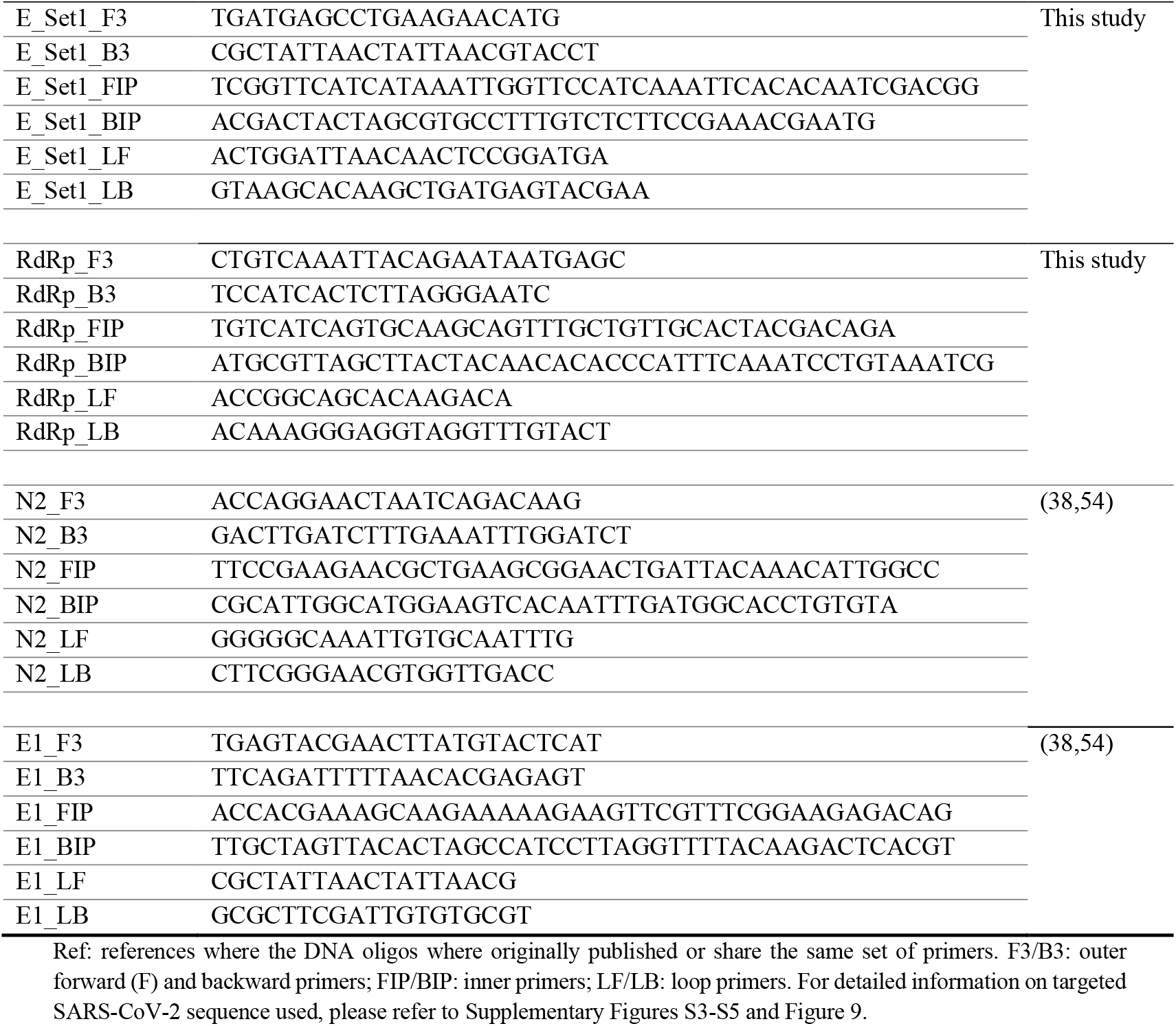
Sets of LAMP oligonucleotides used in this study

### RT-LAMP assays

All mix preparations for RT-LAMP reaction were performed on ice inside a biosafety level 2 cabinet. RT-LAMP reactions were performed according to New England Biolabs (NEB) recommendations, containing the following components: 10 µL of WarmStart^®^ Colorimetric LAMP 2x Master Mix (NEB #M1800 or #M1804, the latter contains dUTP UDG - uracil-DNA-glycosylase - to avoid carryover contamination; composition of both are NEB’s proprietary) – ready to use mixture of WarmStart^®^ *Bst* 2.0 DNA polymerase and WarmStart^®^ RTx (reverse transcriptase for one-step transcription/amplification reaction) in presence of a pH sensor that turns from fuchsia (pink) to yellow in presence of increased proton (acid pH) during DNA polymerization on isothermal amplification; 1.6 µmol/L forward inner/backward inner primers (FIP/BIP); 0.2 µmol/L forward and backward outer primers (F3/B3) and 0.4 µmol/L loop forward and loop backward primers (LF/LB); Ultra-pure™ DNAse/RNase free distilled water (Invitrogen™ #10977015) was added in quantity enough to complete the final volume reaction of 20 µL; isothermal amplification was performed on Veriti™ thermal cycler (Applied Biosystems, Foster City, CA. USA) at 65 ° C for 30 minutes. From clinical samples, we used as input, 1 µL of RNA extracted from nasopharyngeal swab placed on guanidine-containing VTM. When using raw RNA extraction free samples, we initially prepared a 1:10 ultra-pure water diluted clinical sample (1 µL of VTM sample in 9 µL water) and used 1 µL as RT-LAMP reaction input. A similar strategy was applied to SARS-CoV-2 VOC/VOI samples. Positive controls were performed either by RNA extraction from Vero E6-derived inactivated SARS-CoV-2, using synthetic SARS-CoV-2 N gene-harboring plasmid (ECRA Biotech, Campinas, SP, Brasil #EB14-20) or inactivated laboratory-cultured SARS-CoV-2, when aiming the RNA extraction free tests. For optimization purposes, incubation time tested varied from 30 to 50 min. All RT-LAMP reaction products were migrated in 2% agarose gel to confirm: specific amplification in positive reactions and amplicon-free non-template controls. Gel images were taken using the ImageQuant(tm) LAS 4000 with GelRed^®^ (Biotium #41003) as intercalating dye. Non SARS-CoV-2 RNA extracted samples of Influenza A, Influenza B, human respiratory syncytial virus, Dengue, Zika and Chikungunya viruses were also added as 1 µL input.

### Analytical sensitivity

Absolute quantification was performed based on a calibration curve prepared using the standard SARS-CoV-2 E gene-harboring plasmid (2 × 10^5^ copies/µL) (Biogene COVID-19 PCR, Bioclin/Quibasa #K228-1; Lot: 0007); SARS-CoV-2 (2019-nCoV) Charité/Berlin primer probe panel (IDT, #10006804) and the GoTaq^®^ Probe 1-step RT-qPCR System (Promega #A6120), according to manufacturer instructions, as indicated by the United States Centers for Disease Control and Prevention (CDC). Real-time reverse transcriptase-polymerase chain reaction program was performed as follows: 1^st^ stage (x1) 15 min at 45 °C; 2^nd^ stage (x1) 2 min at 95 °C and 3^rd^ stage (x40) 3 sec at 95 °C followed by 30 sec at 55 °C. Linear regression was performed using Prism software, version 9 (GraphPad Software, San Diego, CA, USA) leading to the equation: ***Y*** = -3.6383X + 38.771 and coefficient of correlation ***R***^2^ = 0.9938 (Supplementary Figure S2). Viral RNA either from Vero E6-derived SARS-CoV-2 (SARS-CoV-2 isolate HIAE-02: SARS-CoV2/SP02/human/2020/BRA GenBank accession number MT126808.1) or obtained from clinical nasopharyngeal swabs was quantitated based on the cycle-threshold value for E gene.

## Data Availability

The genomes of SARS-CoV-2 VOI and VOCs generated are deposited on GSAID according to the following accession codes: EPI_ISL_500460, EPI_ISL_500461, EPI_ISL_500865, EPI_ISL_500868, EPI_ISL_500872, EPI_ISL_500477, EPI_ISL_500482, EPI_ISL_1239012, EPI_ISL_1239013, EPI_ISL_1239014, EPI_ISL_1239015, EPI_ISL_1239016; as previously published (Bezerra et al. 2021)
Infect Genet Evol. 2021 May 8;92:104910. doi: 10.1016/j.meegid.2021.104910.

## ACKNOWLEDGMENTS

We thank Professor Édison L. Durigon, Laboratório de Virologia Clínica e Molecular, ICB/USP, São Paulo, Brazil, who kindly provided inactivated samples of SARS-CoV-2 isolate HIAE-02: SARS-CoV2/SP02/human/2020/BRA (GenBank accession number MT126808.1). We thank Dr. Luciano Moreira, Instituto René Rachou – Fiocruz Minas, for providing the first colorimetric LAMP buffers during the testing and optimization phase. We also thank Dr. Marluce Aparecida Assunção Oliveira – Former Director of Instituto Octávio Magalhães, Fundação Ezequiel Dias, Laboratório Central de Saúde Pública de Minas Gerais – LACEN-MG; and Dr. Marcos Vinicius Ferreira da Silva, head of virology service at LACEN-MG, for providing samples of Influenza A and B viruses and human respiratory syncytial virus (hRSV). We are grateful to to Cristiane P. Gomes and Patrícia P. N. Miranda for resources management and excellent technical assistance. This work was funded by: Fundação de Amparo à Pesquisa do Estado de Minas Gerais – Fapemig, grant number #APQ-00485-20, to RLMN; grant number #APQ-00262-20 to F.D.C. and Fundação Oswaldo Cruz – Inova Fiocruz Program – Innovative Products (grant number VPPIS-004-FIO-18-51) to RLMN; Innovative Products to face COVID-19 pandemics (grant number VPPIS-005-FIO-20-2-45) to PAA and from the MCTI – Brazilian Ministry of Science, Technology and Innovation, through the “*Rede Virus*” initiative to P.A.A (grant number – FINEP 01.20.0005.00). E.G.O. received a Master’s fellowship from the Vice presidency of Education, Formation and communication VPEIC – Fundação Oswaldo Cruz. IAB and APMFL received post-doctoral research fellowships from Inova Fiocruz Program – Fundação Oswaldo Cruz. ABG holds a science, technology and innovation development scholarship from Fapemig (BDCTI-I). LAT is a post-doctoral fellow from MCTI (CNPq – DTI-A). R.L.M.N., S.M.R.T., G.W. and G.L.W. are CNPq Research Fellows (#310640/2017-2; #302961/2017-8; #303902/2019-1; #307538/2019-2). P.M holds a scholarship from Coordenação de Aperfeiçoamento de Nível Superior (CAPES/Ação Emergencial).The funders had no role in the study design, data collection and analysis, decision to publish or manuscript preparation. P.A.A, G.L.W and R.L.M.N. are part of Fiocruz COVID-19 Genomic Surveillance Network

