## Supplementary material for "Clinical validation of colorimetric RT-LAMP, a fast, highly sensitive and specific COVID-19 molecular diagnostic tool that is robust to detect SARS-CoV-2 variants of concern"

**Table S1 – SARS-CoV-2-targeting primers showing their melting reference temperatures and coverages.** Some N and E genes-targeting primers presented cross-reactivity with SARS-CoV genomes.

| Primer ID | $T_m$ (°C) | Coverage (%) | Number of crossing genomes | |
|---|---|---|---|---|
| | | SARS-CoV-2 | SARS-CoV | non-SARS-CoV |
| SARS-CoV-2_N_Set1_B1c | 69.2 | 98.0 | - | - |
| SARS-CoV-2_N_Set1_B2 | 60.6 | 99.7 | - | - |
| SARS-CoV-2_N_Set1_B3 | 62.5 | 99.6 | - | - |
| SARS-CoV-2_N_Set1_F1c | 68.1 | 99.6 | - | - |
| SARS-CoV-2_N_Set1_F2 | 61.9 | 98.1 | - | - |
| SARS-CoV-2_N_Set1_F3 | 61.6 | 99.5 | 9 | - |
| SARS-CoV-2_N_Set1_LB | 64.2 | 99.5 | 9 | - |
| SARS-CoV-2_N_Set1_LF | 69.3 | 99.7 | - | - |
| SARS-CoV-2_N_Set2_B1c | 70.1 | 99.8 | - | - |
| SARS-CoV-2_N_Set2_B2 | 62.9 | 99.6 | - | - |

| | | | | |
|---|---|---|---|---|
| SARS-CoV-2_N_Set2_B3 | 64.6 | 99.3 | - | - |
| SARS-CoV-2_N_Set2_F1c | 69.4 | 99.7 | 10 | - |
| SARS-CoV-2_N_Set2_F2 | 62.8 | 97.1 | - | - |
| SARS-CoV-2_N_Set2_F3 | 61.7 | 99.7 | - | - |
| SARS-CoV-2_N_Set2_LB | 67.6 | 99.7 | 10 | - |
| SARS-CoV-2_N_Set2_LF | 66.4 | 99.6 | 1 | - |
| SARS-CoV-2_E_Set1_B1c | 62.5 | 99.6 | 9 | - |
| SARS-CoV-2_E_Set1_B2 | 59.7 | 98.7 | - | - |
| SARS-CoV-2_E_Set1_B3 | 59.5 | 99.7 | 10 | - |
| SARS-CoV-2_E_Set1_F1c | 68.6 | 99.9 | 9 | - |
| SARS-CoV-2_E_Set1_F2 | 60.8 | 95.0 | - | - |
| SARS-CoV-2_E_Set1_F3 | 61.3 | 99.6 | - | - |
| SARS-CoV-2_E_Set1_LB | 70.6 | 99.3 | - | - |
| SARS-CoV-2_E_Set1_LF | 66.9 | 99.6 | - | - |
| SARS-CoV-2_RdRp_Set1_B1c | 63.4 | 99.6 | - | - |
| SARS-CoV-2_RdRp_Set1_B2 | 64.6 | 99.5 | - | - |
| SARS-CoV-2_RdRp_Set1_B3 | 60.2 | 99.4 | - | - |
| SARS-CoV-2_RdRp_Set1_F1c | 65.7 | 99.9 | - | - |
| SARS-CoV-2_RdRp_Set1_F2 | 56.6 | 99.9 | - | - |
| SARS-CoV-2_RdRp_Set1_F3 | 62.4 | 99.7 | - | - |
| SARS-CoV-2_RdRp_Set1_LB | 63.8 | 99.3 | - | - |
| SARS-CoV-2_RdRp_Set1_LF | 60.2 | 99.9 | - | - |

The RT-LAMP primer sets were aligned to 3364 SARS-CoV-2 genomes and 323 genomes belonging to other coronaviruses being 10 SARS-CoV-related and 313 non-SARS-CoV ones. Melting temperatures were calculated based on mesoscopic model considering up to three mismatches (1). FIP = F1c+F2; BIP = B1c+B2. N: SARS-CoV-2 Nucleocapsid protein coding sequence; E: SARS-CoV-2 Envelope protein coding sequence and RdRp: SARS-CoV-2 RNA-dependent RNA polymerase coding sequence

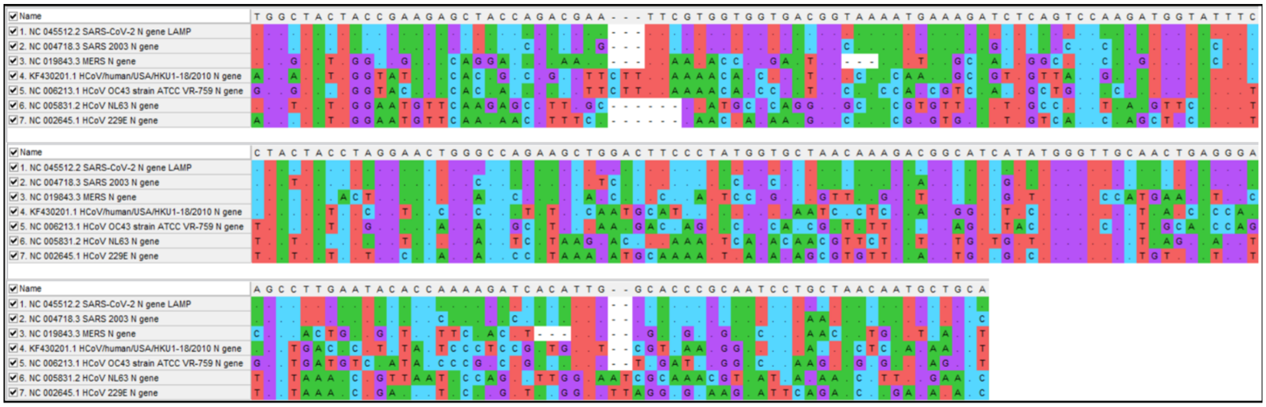

**Figure S1 – Multiple sequence alignment of partial SARS-CoV-2 N gene compared to other human coronaviruses.**

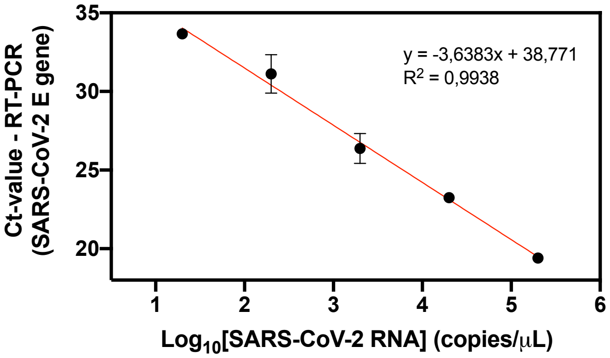


**Figure S2 – Primer efficiency or calibration curve used for analytical sensitivity quantification**. RT-qPCR was performed using SARS-CoV-2 E gene as target (Charité/Berlin protocol) using the SARS-CoV-2 E gene-harboring plasmid (2 x $10^5$ copies/µL) (Biogene COVID-19 PCR, Bioclin/Quibasa #K228-1; Lot: 0007). Linear regression was used to calculate the limit of detection.

**Table S2 –** Comparative colorimetric RT-LAMP using two distinct set of primers targeting N gene on low viral load, nasopharyngeal RNA extracted samples.

| RT-qPCR Ct value (E gene) | RT-qPCR Status | RT-LAMP - SARS-CoV-2 | | |
|---|---|---|---|---|
| | | N gene_Set1 | N gene_Set2 | Multiplex N gene_Set1/Set2 |
| 31.85 | Positive | Negative | ND | Positive |
| 32.07 | Positive | Positive | Positive | Positive |
| 32.1 | Positive | Negative | Positive | Positive |
| 32.56 | Positive | Negative | ND | Positive |
| 32.59 | Positive | Negative | Positive | Positive |
| 32.98 | Positive | Positive | Positive | Positive |
| 33.18 | Positive | Negative | ND | Negative |
| 33.35 | Positive | Negative | Positive | Positive |
| 33.6 | Positive | Negative | Positive | Positive |
| 34.31 | Positive | Positive | Positive | Positive |
| 35.2 | Positive | Negative | ND | Positive |
| 36.19 | Positive | Positive | Positive | Positive |

**Table S3 –** Clinical SARS-CoV-2 sequenced samples isolated in Pernambuco State, Brazilian Northeast, depicting the lineage or variant identified.

| Lineage/Variant | Isolated at (City, State) |
|---|---|
| P.1 | Jaboatão dos Guararapes, Pernambuco |
| P.1 | Recife, Pernambuco |
| P.2 | Poção, Pernambuco |
| P.2 | Recife, Pernambuco |
| B.1.1.374 | Cabo de Santo Agostinho, Pernambuco |
| B.1.1 | Recife, Pernambuco |
| B.1.1.28 | Igarassu, Pernambuco |
| B.1.1.371 | Recife, Pernambuco |

SARS-CoV-2 RNA was reversely transcribed into cDNA and sequenced by NGS (Illumina) according to the previous work by (2). Variant of concern/interest monitoring are being currently monitored by Sanger sequencing of specific polymorphic regions, such as Spike-RBD coding portion (K417N/T, E484K, N501Y, A570D) (3). SARS-CoV-2 lineages and variants nomenclature follows the PANGO lineages proposal (https://cov-lineages.org/)(4). SARS-CoV-2 variant genomes were deposited on GISAID according to the following accession IDs: EPI_ISL_2221860, EPI_ISL_2221850, EPI_ISL_2221873, EPI_ISL_2221890, EPI_ISL_2221902, EPI_ISL_2221885, EPI_ISL_2221844, EPI_ISL_2221866

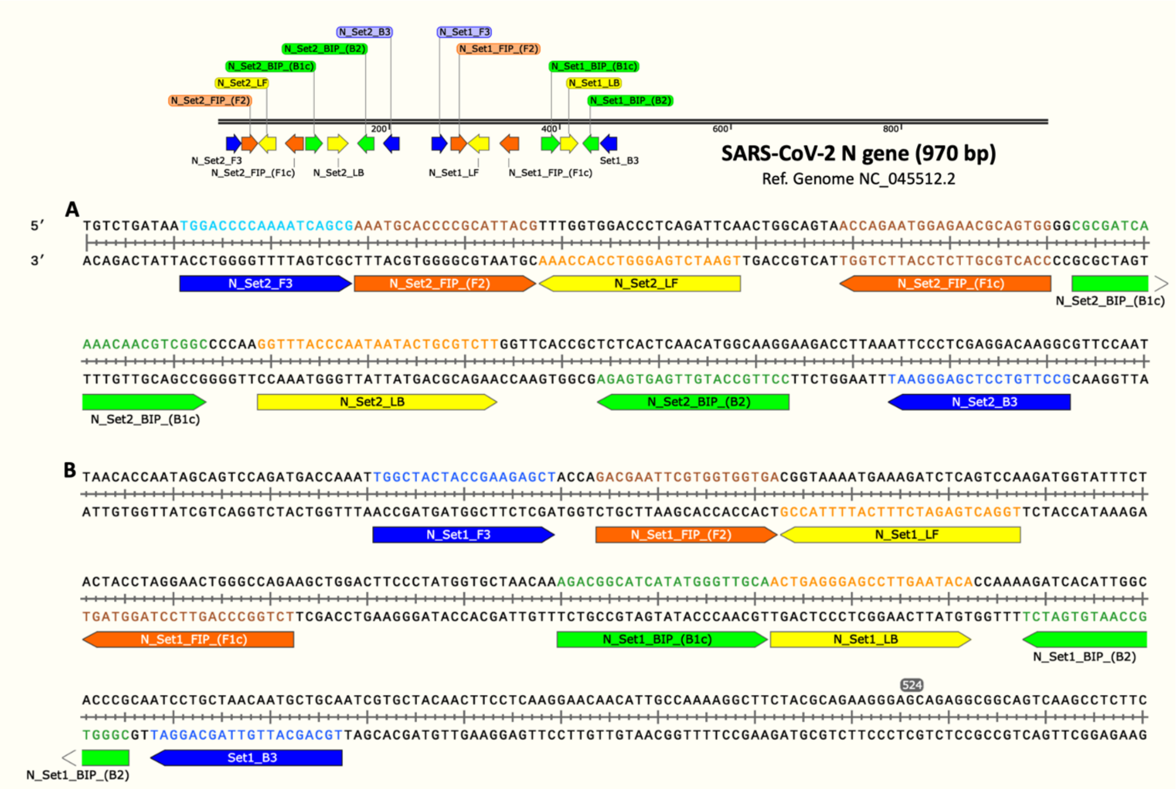


**Figure S3** – SARS-CoV-2 specific N gene set of LAMP primers aligned to their respective target region for Set1 (A) and Set2 (B). The upper scheme represents the zoomed out region on SARS-CoV-2 reference genome NC_045512.2. Primers were designed using PrimerExplorer V5 or NEB LAMP primer designer softwares. F3/B3: outer primers; FIP/BIP: inner primers and LF/LB: loop primers. Created on SnapGene Viewer

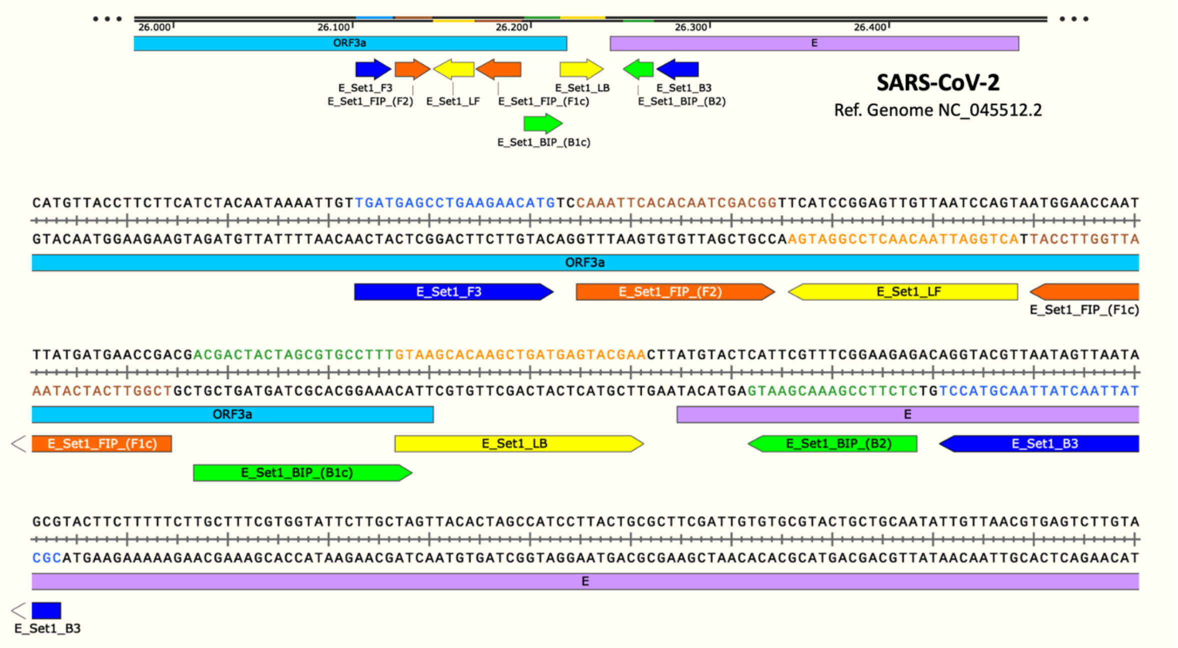


**Figure S4** – SARS-CoV-2 E gene set of LAMP primers aligned to their respective target region, designed based on NC_045512.2 reference genome. F3/B3: outer primers; FIP/BIP: inner primers and LF/LB: loop primers. E: envelope protein-coding sequence. Created on SnapGene Viewer

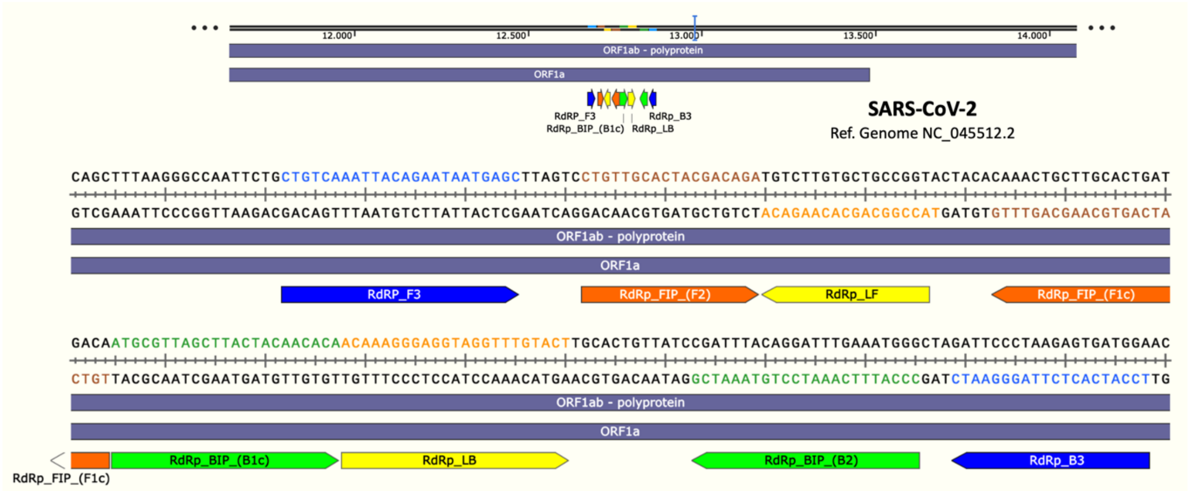


**Figure S5** – SARS-CoV-2 RdRp gene set of LAMP primers aligned to their respective target region, designed based on NC_045512.2 reference genome. F3/B3: outer primers; FIP/BIP: inner primers and LF/LB: loop primers. RdRp: viral RNA-dependent RNA polymerase-coding sequence. Created on SnapGene Viewer
